# A Fast Nonparametric Sampling (NPS) Method for Time-to-Event in Individual-Level Simulation Models

**DOI:** 10.1101/2024.04.05.24305356

**Authors:** David U. Garibay-Treviño, Hawre Jalal, Fernando Alarid-Escudero

**Author notes:** **Corresponding Author:** David U. Garibay-Treviño, M.P.P., 600 Peter Morand, 301, Ottawa, Ontario, Canada. K1N 6N5.

## Abstract

**Purpose:** Individual-level simulation models often require sampling times to events, however efficient parametric distributions for many processes may often not exist. For example, time to death from life tables cannot be accurately sampled from existing parametric distributions. We propose an efficient nonparametric method to sample times to events that does not require any parametric assumption on the hazards.

**Methods:** We developed a nonparametric sampling (NPS) approach that simultaneously draws multiple time-to-event samples from a categorical distribution. This approach can be applied to univariate and multivariate processes. We discretize the entire period into equal-length time intervals and then derived the interval-specific probabilities. The times to events can then be used directly in individual-level simulation models. We compared the accuracy of our approach in sampling time-to-events from common parametric distributions, including exponential, gamma, and Gompertz. In addition, we evaluated the method’s performance in sampling age to death from US life tables and sampling times to events from parametric baseline hazards with time-dependent covariates.

**Results:** The NPS method estimated similar expected times to events from 1 million draws for the three parametric distributions, 100,000 draws for the homogenous cohort, 200,000 draws from the heterogeneous cohort, and 1 million draws for the parametric distributions with time-varying covariates, all in less than a second.

**Conclusion:** Our method produces accurate and computationally efficient samples for time-to-events from hazards without requiring parametric assumptions.

## Introduction

Discrete-event simulation (DES) models simulate processes as discrete sequences of events that occur over time.^1^ These models rely on sampling the time of different events. For example, if events have a constant rate or hazard of occurrence, the time of their occurrence can be sampled from an exponential distribution. In DES models, time-to-event data following a nonconstant hazard could be sampled from parametric distributions.^2^ However, some events cannot be easily described by parametric distributions. For example, life tables, or events following hazards that are a function of time-varying covariates, such as smoking histories or tumor size, do not always follow standard parametric distributions. An alternative is to use a nonhomogeneous Poisson point process (NHPPP), which assumes that the rate of events follows a Poisson process that can vary over time.^3^ There are different implementations of algorithms for sampling from NHPPP, which require either numerical integration or rejection sampling.^4^

In this brief report, we propose a nonparametric sampling (NSP) implementation of NHPPP that is both generalizable and computationally efficient. The method assumes that time to event is drawn from a nonparametric categorical distribution. We illustrate the NPS method using 5 examples highlighting its accuracy, flexibility and computational efficiency. Additionally, we provide an open-source implementation in R and Python to facilitate wider adoption.

### Constructing the categorical distribution

The steps to implement the NPS method are described in Box 1 and shown in Figure 1. In summary, the approach involves six steps: First, obtaining the discrete-time hazard cumulative distribution function, *F*_*t*_. This can be obtained from nonparametric estimation methods, such as the life table, actuarial or Kaplan-Meier methods.^5^ Second, obtaining the interval-specific sample probability, which is the probability mass function, *p*_*t*_, derived from *F*_*t*_. Third, sampling the times to events employing a categorical distribution, using the interval-specific probabilities, and defining each time interval as a category. And lastly, approximating time to event in continuous time. If *F*_*t*_ is not readily available, it can be derived from either the discrete-time cumulative hazard, *H*_*t*_, (e.g., obtained from a Nelson-Aalen estimator^6^), or the interval-specific discrete-time hazard, *h*_*t*_ (e.g., obtained from a life table or actuarial estimation method). If instead, the cumulative hazard is available in continuous time, *H*_*t*_, then *h*_*t*_ can be derived within a time interval Δ*t* from *H*(*t* + Δ*t*) − *H*(*t*). Below we provide further details on these steps.

**Figure 1.**
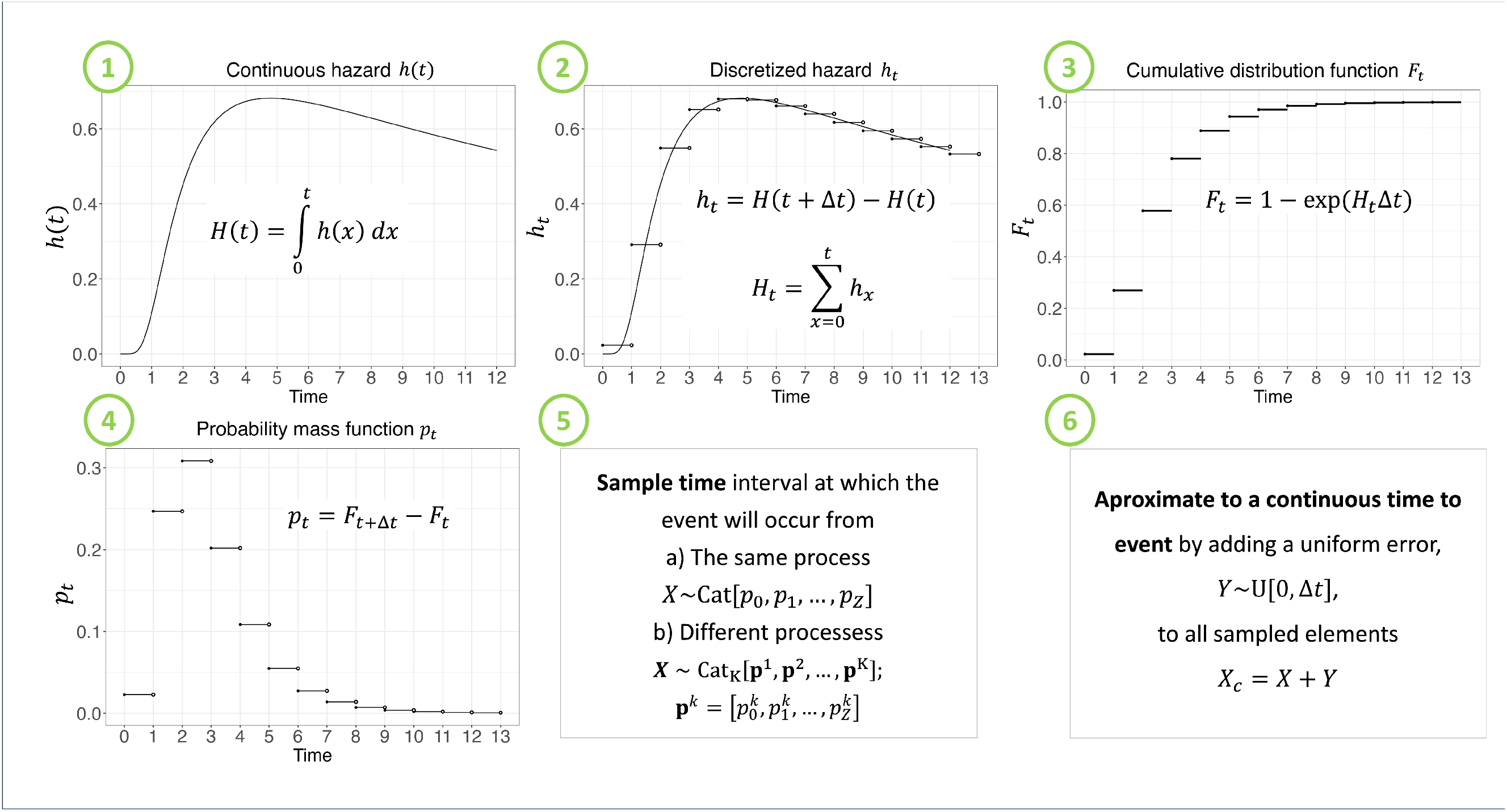
Steps to sample time-to-events using a nonparametric sampling (NPS) approach.

Let *t* ∈ *T* be the time-to-event following a piecewise constant hazard *h*_*t*_ within a time interval *Δt*, where *T* is a random variable, *t* = 0,…, *Z*, and *Z* is the last time interval by which the event can occur. Thus, the cumulative hazard function at time *t, H*_*t*_, is obtained from

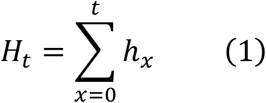

The cumulative distribution function (CDF) of *T* at time *t, F*_*t*_, is

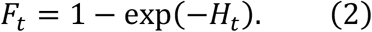

If the hazard is given in a different scale from the one the analyst is interested in, it can be transformed to the desired scale by multiplying the hazard *h*_*t*_ by *Δt*, where *Δt* represents the ratio of the given scale to the desired scale.^5^ For example, if the hazards are on a yearly scale and we want to sample monthly time-to-event data, we use, 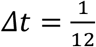, and when the samples are in years, we use *Δt* = 1. This scale transformation assumes that the hazard is constant within the interval.

We derive the probability of an event happening within the *t*-th interval [*t, t* + *Δt*) by the difference in the CDF in Equation 2 as

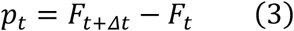

To conduct a non-parametric sampling (NPS) of the time interval at which the event can occur, we define *X* as the time interval at which the event can occur and assume it follows a categorical distribution where each time interval is considered a category. Thus,

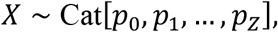

with a probability mass function *f*(*X* = *x*|**p**) = *p*_*t*_, where **p** = (*p*_0_, *p*_1_,…, *p*_)_), *p*_*t*_ *≥* 0 is the probability of the event occurring at the *t*-th time interval [*t, t* + *Δt*) and 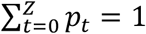. Most statistical software provide built-in functions to sample from a categorical distribution. For example, in `R` is the `sample` function, and in `Python` is the `numpy.random.choice` function.

### Multivariate categorical distribution

We expand the previous approach to sample values for multiple random variables simultaneously by defining a multivariate categorical distribution as

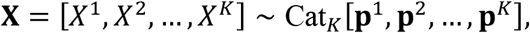

where *X^k^* = *x^k^* is the *k*-th random variable with a vector of probabilities *p^k^* of having the events in each of the time intervals defined as

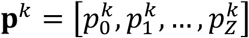

Common statistical software has no built-in functions to sample from a multivariate categorical distribution. However, we provide the code of the multivariate categorical distribution in `R` and `Python` in the Supplementary material.

### Approximating continuous time-to-event

An approximation error occurs when approximating continuous time-to-event by using a discrete-time approach.^7–9^ Since the NPS samples for the exact time categories that were initially defined while dividing the time interval, the method does not contemplate the possibility of events happening in between any two categories. This generates a systematic bias, which could be reduced by adding a random variable *Y* ∼ *U*[0, *Δt*] to *X*, assuming that the time to event within each *Δt* interval is equally likely to occur within the interval, which is consistent with a piecewise constant hazard model. Thus, the random variable of the time to event with the correction, is *X*, = *X* + *Y*. The steps to sample time to event using an NPS are shown in Box 1 and illustrated in Figure 1. Adding a random uniform value to each time to event sampled from the categorical NPS method and calculating the expected value across all the samples is equivalent to adding a *Δt*/2 value to the expected time to event, which is a half-cycle correction (HCC) often used in discrete-time cohort state transition models^10^. If there is interest only in the expected value of the time to events, analysts could add a *Δt*/2 value to the expected value of the time to event obtained from the categorical NPS method without correction.

### Accounting for covariates

Hazards could be a function of either of time-independent covariates, such as sex, race, or birth cohort, or time-dependent covariates, such as smoking histories, exposure to environmental risk factors or tumor size. In this section, we demonstrate the use the NPS method to sample times to events from hazards as functions of time-independent and time-dependent covariates.

#### Time-independent covariates

Let the *i*-th individual time to an event *T*_*i*_ follow a time-dependent hazard, *h*_*i*_(*t*) = *f*_*i*_(*x*_*i*_; *β*), over a time interval [0, *Z*], as a function of a time-independent covariate, *x*_*i*_, that can take any functional form and vary between individuals, and a set of coefficients *β*. We assume a proportional hazards approach of the effect of the covariates on the hazard to demonstrate how to sample time to events the NPS method. That is, 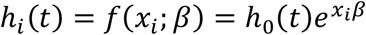, where *h*_0_(*t*) is the time-dependent baseline hazard, *x*_*i*_ is the covariate for the *i*-th individual and *β* is the log-hazard ratio of the proportional effect of the covariate *x*_*i*_ on *h*_0_(*t*).

#### Time-dependent covariates

We now consider that the covariate can vary over time *x*_*i*_(*t*) and can take any functional form, which results in a time-varying hazard *h*_*i*_(*t*) = *f*_*i*_(*x*_*i*_(*t*); *β*), over a time interval [0, *Z*]. The time-dependent covariate could be the same across all individuals (e.g., all experiencing the same mean tumor growth over time) or vary by individuals (e.g., everyone having their own smoking history). To use the NPS method to sample from hazards with time-varying covariates, we generate or pre-specify the time-dependent covariate and compute the corresponding hazard. For example, Figure 2 shows a time-dependent Weibull hazard. We use the multivariate categorical distribution to sample time to events for multiple individuals with different covariate paths.

**Figure 2.**
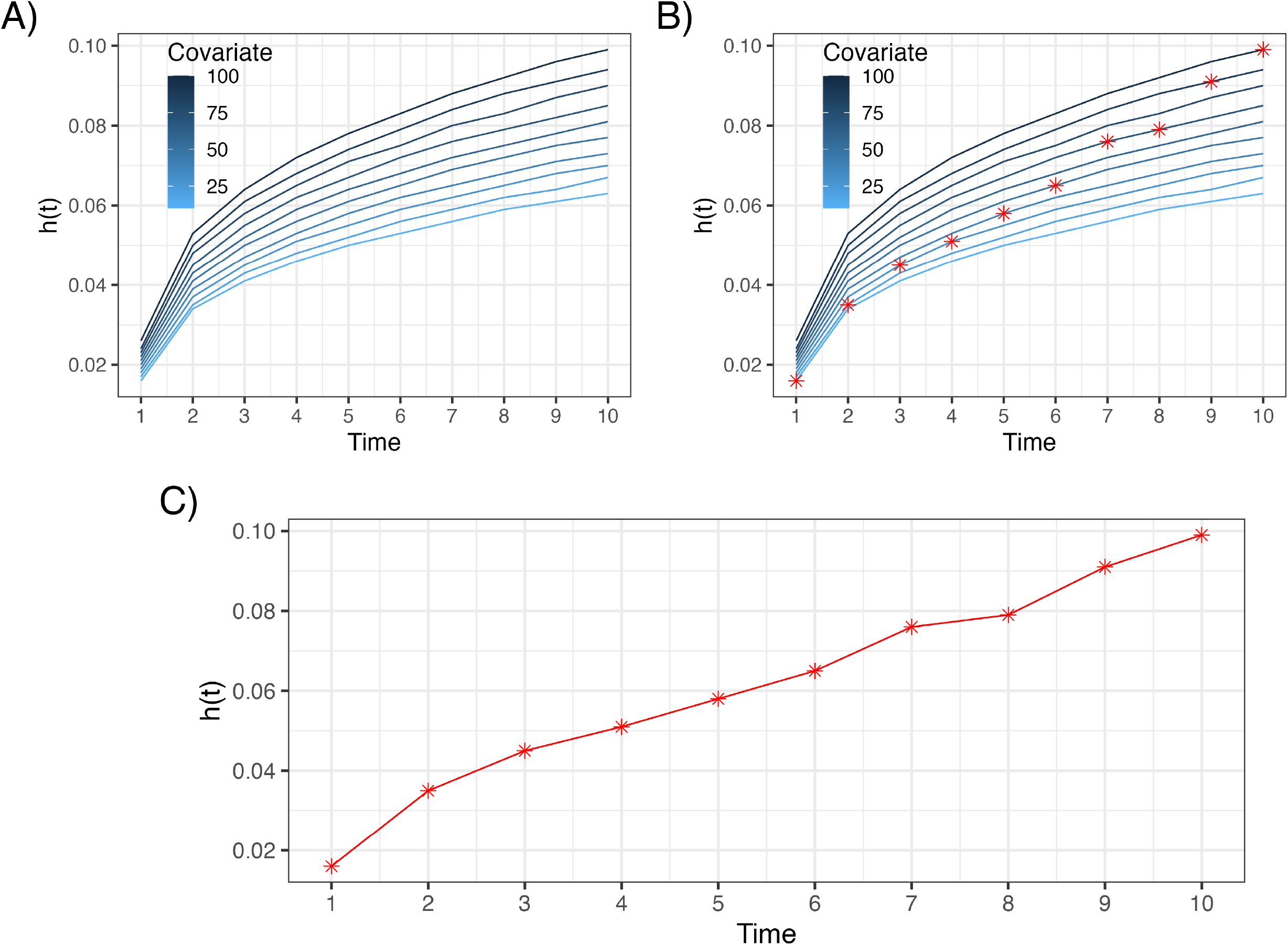
A) Time-dependent hazard, *h*(*t*), for different values of a covariate; B) example of a covariate path; C) Corresponding path of the *h*(*t*).

### Examples

Below, we provide 5 examples to illustrate the implementation of the NPS method for different processes. The R code for these examples and the function of the multivariate categorical distribution is provided in a GitHub repository (https://github.com/DARTH-git/NPS_time_to_event).

#### Example 1: Time to event from parametric hazards

We used the NPS method for drawing times to events from various commonly used parametric distributions, such as exponential, gamma, and log-normal. We derived the piece-wise constant hazard, *h*_*t*_, as described in step 1 in Figure 1 and Box 1 and applied Equation 2 and Equation 3. We sampled 10,000 times to event, computed the mean across all samples, and repeated this 1,000 times to compute the overall mean across all simulations. We then compared the expected time to event obtained from our method, with and without the approximation to continuous-time interval, to the analytic expected time from the parametric distributions. While the expected times to event coming from the exponential, gamma, and log-normal distributions, using the NPS method and accounting for continuous time, were 10, 39.98, and 33.49, their analytical values were 10, 40, and 33.49, respectively. We also computed the mean execution time, and their 95% interquantile range (IQR, see Table 1), from 100 iterations using a computer with 2.3GHz Quad-Core Intel Core i7 with 32GB memory. The mean execution time for the exponential, gamma and log-normal distributions were 0.49, 0.61 and 0.54 milliseconds, respectively.

**Table 1:** Comparison of expected time to events and mean sampling time, in milliseconds, from 100 iterations of N samples each between the non-parametric sampling (NPS) method and parametric distributions or life table estimates. NPS-U: Non-parametric sampling uncorrected; NPS-C: Non-parametric sampling corrected by adding a uniformly distributed random number; N/A: Not applicable.

| Distribution | NPS-U | NPS-C | Analytic solution | Parametric sampling |
| --- | --- | --- | --- | --- |
| **Exponential (rate = 0.1; N = 10,000)** |  |  |  |  |
| Expected value | 9.51 | 10.00 | 10.00 | 10.00 |
| Mean execution time (95% IQR) | 0.35[0.31, 0.48] | 0.49[0.43, 0.66] |  | 0.45[0.35, 0.69] |
| **Gamma (rate = 0.1, shape = 4; N = 10,000)** |  |  |  |  |
| Expected value | 39.47 | 39.98 | 40.00 | 40.00 |
| Mean execution time (95% IQR) | 0.49[0.39, 0.68] | 0.61[0.50, 0.85] |  | 0.88[0.68, 3.12] |
| **Log-normal ($\mu = 3.5$, $\sigma = 0.15$; N = 10,000)** |  |  |  |  |
| Expected value | 32.99 | 33.49 | 33.49 | 33.49 |
| Mean execution time (95% IQR) | 0.34[0.30, 0.49] | 0.54[0.42, 0.78] |  | 0.64[0.54, 0.97] |
| **Life tables - Homogeneous cohort; N = 100,000** |  |  |  |  |
| Expected value | 78.04 | 78.53 | 78.37 | N/A |
| Mean execution time (95% IQR) | 3.63[3.23, 4.05] | 5.15[4.38, 5.36] |  |  |
| **Life tables - Heterogeneous cohort; N = 200,000** |  |  |  |  |
| Expected value (females) | 80.32 | 80.93 | 80.76 | N/A |
| Expected value (males) | 75.71 | 76.22 | 75.93 |  |
| Mean execution time (95% IQR) | 254.36[217.37, 394.44] | 255.30[219.65, 409.17] |  |  |

#### Example 2: Sampling age to death from a homogeneous cohort

We sampled the age to death for 100,000 individuals in a hypothetical cohort from the US population in 2015.^11^ We estimated the life expectancy by taking the average across the 100,000 samples with the continuous-time approximation. The probability mass function (PMF) for the age to death obtained from the NPS methods closely follows the PMF from the life table (Figure 3). The estimated life expectancy from the NPS method is 78.53 years, which is close to the life expectancy obtained from the life tables of 78.37 years. The mean execution time, repeating the sampling process 100 times is 5.15 milliseconds (Table 1).

**Figure 3.**
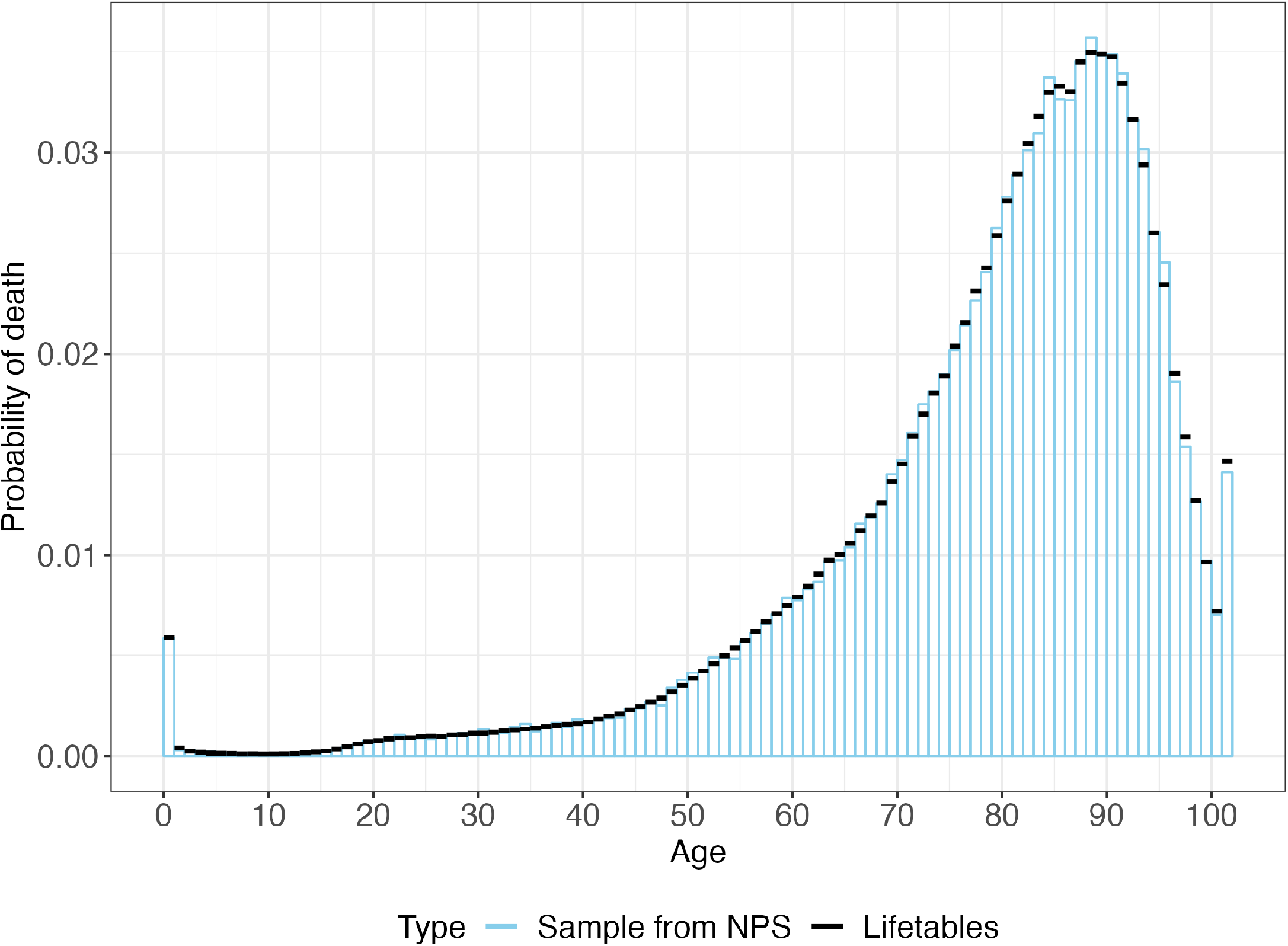
Probability mass function (PMF) of dying within a year of age in the total U.S. population in 2015.

#### Example 3: Drawing age to death from a heterogeneous cohort

We used the multivariate categorical distribution to simultaneously sample ages to death for 100,000 males and females from sex-specific life tables for the U.S. population in 2015, with the continuous-time approximation defined above. The sex-specific PMF from the NPS method and the exact PMF from life tables are shown in Figure 4. The NPS method estimated a life expectancy of 76.22 and 80.93 years for males and females, respectively. The life expectancy obtained from the life tables was 75.93 and 80.76 years for males and females, respectively. The mean execution time, repeating the sampling process 100 times is 255.30 milliseconds (Table 1).

**Figure 4.**
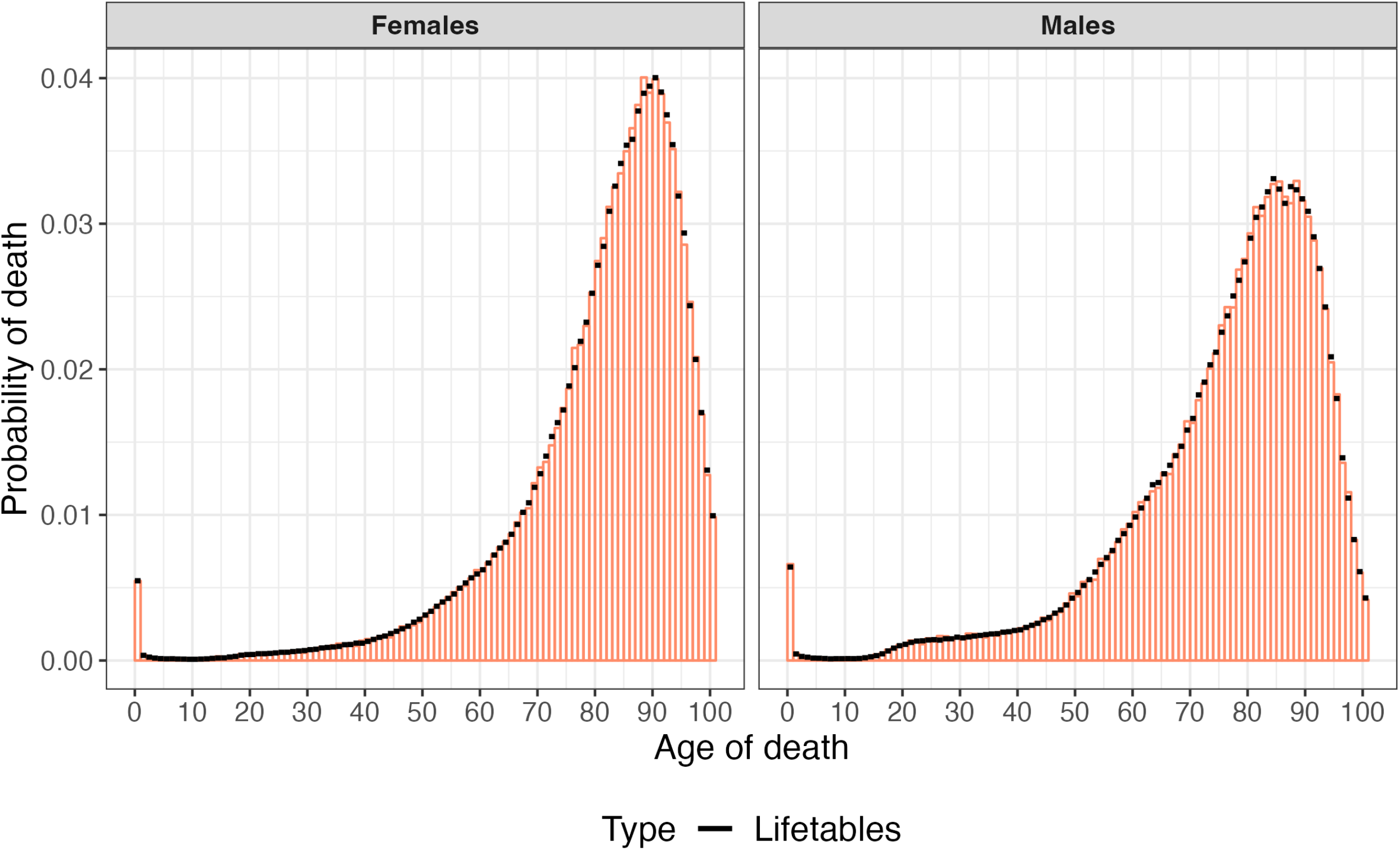
PMF of dying within a year of age by sex, U.S. population in 2015.

#### Example 4: Drawing time to event from hazards with time-dependent covariates

We used a proportional hazard setup with a time-dependent covariate that increases linearly over time, *x*_*i*_(*t*) = *α*_0_ + *α*_1_*t*, obtaining 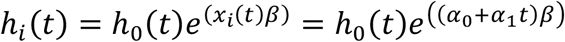. We compared the accuracy of the method in sampling time-to-events from parametric exponential (rate = 0.1) and Gompertz (shape = 0.1, scale = 0.001) baseline hazards, considering a linear time-varying covariate (*α*_0_ = 0, and *α*_1_ = 1) with a log-hazard ratio (*β* = 1.02) against those obtained using direct sampling (DS) from the inverse cumulative density functions obtained analytically.^12,13^

The NPS method produced similar expected time to events for the two distributions compared to the DS method, from 1 million draws: exponential (8.61 NPS vs. 8.52 DS), Gompertz (35.98 NPS vs. 35.48 DS), and Weibull (8.79 NPS vs. 8.02, DS). Their mean execution time in milliseconds, repeating the sampling process 100 times, is 38.28, 51.92, and 48.44, respectively.

#### Example 5: Drawing time to event from hazards with time-dependent covariates following random paths

We specify a time-varying covariate *x*_*i*_(*t*) = *α*_0_ + *α*_1_*y*_*i*_(*t*) assuming *y*_*i*_(*t*) follows a Gaussian random walking process *y*_*i*_(*t*) = *y*_*i*_(*t* − 1) + *ϵ*_*i*_, where *ϵ*_*i*_ ∼ *Normal*(*μ* = 0, *σ* = 0.5) and generated 1,000 random paths over 100 years (Figure 5). We assume a Weibull baseline hazard, *h*_0_(*t*) = *Weibull*(*shape* = 1.3, *scale* = 30.1), obtaining 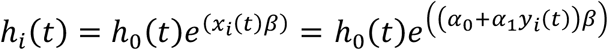. We used the multivariate categorical distribution to sample times to events from the individual-level Gaussian random walk processes and estimated an expected time to event of 27.88 years. Repeating the sampling process 100 times, the average sampling time was 4.27 milliseconds.

**Figure 5.**
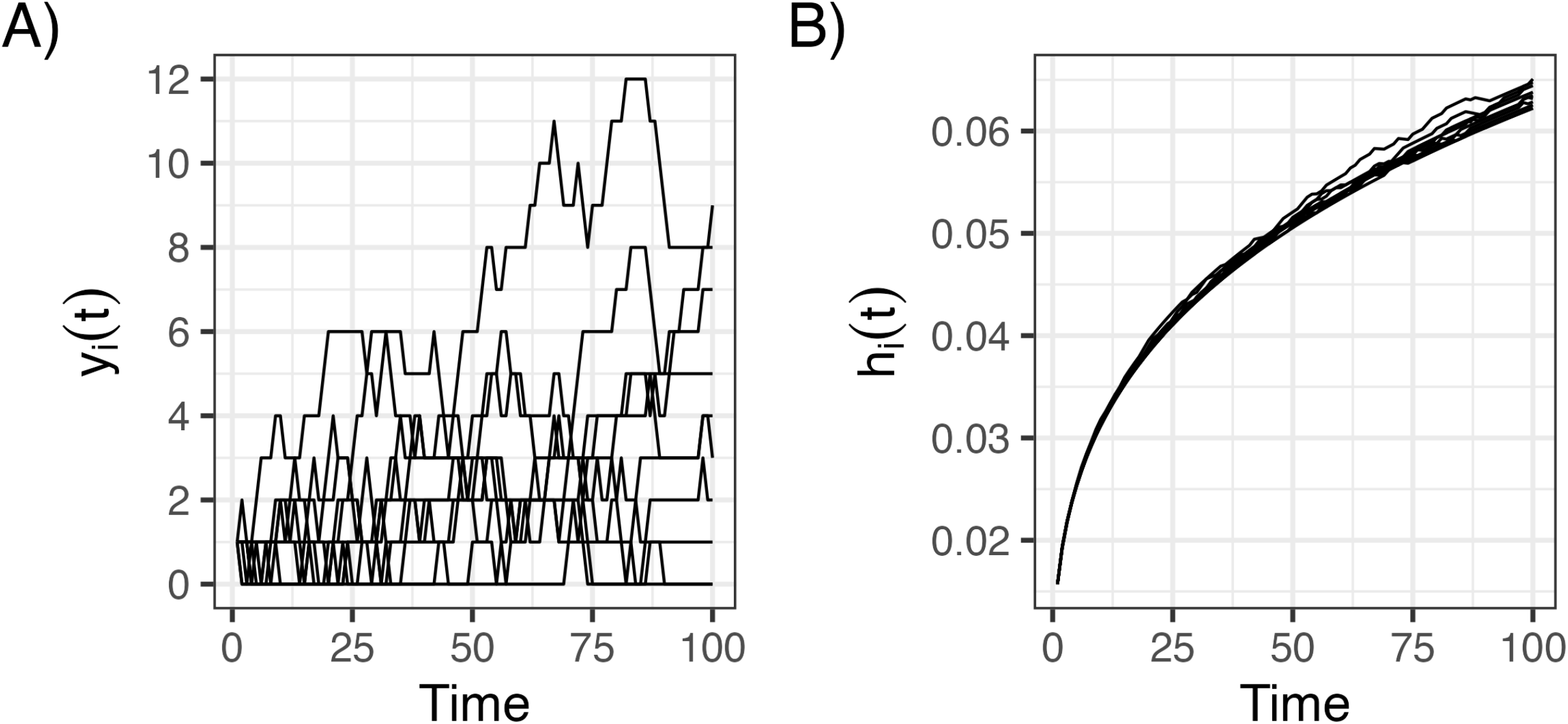
A) Individual-specific trajectories. B) Individual-specific time-dependent hazards. Sample of 10 individuals

## Discussion

We developed a nonparametric approach method to sampling times to events with high computation efficiency. The NPS method uses a categorical distribution, which discretizes the hazard of events over a fixed and finite time period, assuming a piecewise hazard. We illustrated the NPS method with five examples that show common situations encountered when building DES models and provided their mean execution times. NPS can be used to sample the age of death from age-, sex-, race-, and year-specific life tables, and or times to smoking initiation or cessation from smoking histories^14^. It can also be used to sample times to events with hazards that are functions of either time-independent or time-dependent covariates.

The proposed NPS method works similarly to previous methods when sampling ages of death from a life table for a specific group (e.g., white females born in 1980 in the US) using a piecewise-constant exponential distribution^15^. However, a strength of the proposed method is the use of multivariate categorical sampling, which extends the NPS method to simultaneously sample multiple ages of death from multiple life tables for different groups.

The NPS method accurately approximates the expected time to events from parametric distributions and can generate times to events from hazards for which no parametric distributions can be accurately fitted, such as time-varying hazards described by time-varying covariates. Once the probability distributions are derived from the observed hazards, the sampling process is computationally efficient and can be easily repeated multiple times. This approach can be very useful for individual-level models that require sample times to events following processes that could not be appropriately addressed using parametric distributions.

Our approach does not provide criteria to determine the optimal time interval length and it is up to the user to define it. This may pose a limitation because selecting an excessively wide interval can result in distributions that do not resemble the observed hazard, such as those with extremely swift changes in their levels. However, this is a focus for future research. Additionally, since this method utilizes a nonparametric categorical distribution, a sufficient number of samples must be drawn to obtain unbiased estimates. Our method assumes that the analyst is interested in sampling time to events from the mean process. However, if the analyst is interested in propagating the uncertainty of the estimated time-to-event process and has access to the mechanism generating the uncertainty of the estimation of the average process, the user can sample multiple hazards from this mechanism and apply our method to each of the sampled hazards.

We proposed a method that can efficiently sample times to event from any time-to-event process from its hazard, survival, or cumulative distribution function over time. Moreover, this method can simultaneously sample from multiple different hazards with the multivariate categorical distribution, which we provide as R and Python functions in the Supplementary material.

## Data Availability

The code and data used to generate all the examples, are available online using the data availability link.

https://github.com/DARTH-git/NPS_time_to_event

## Acknowledgements

We thank Rowan Iskandar for his valuable contributions to the code for the multivariate categorical sampling. We thank Karen Kuntz, Thomas Trikalinos and Yuliia Sereda for providing feedback on an earlier version of this manuscript.

## Boxes

**Box 1:**
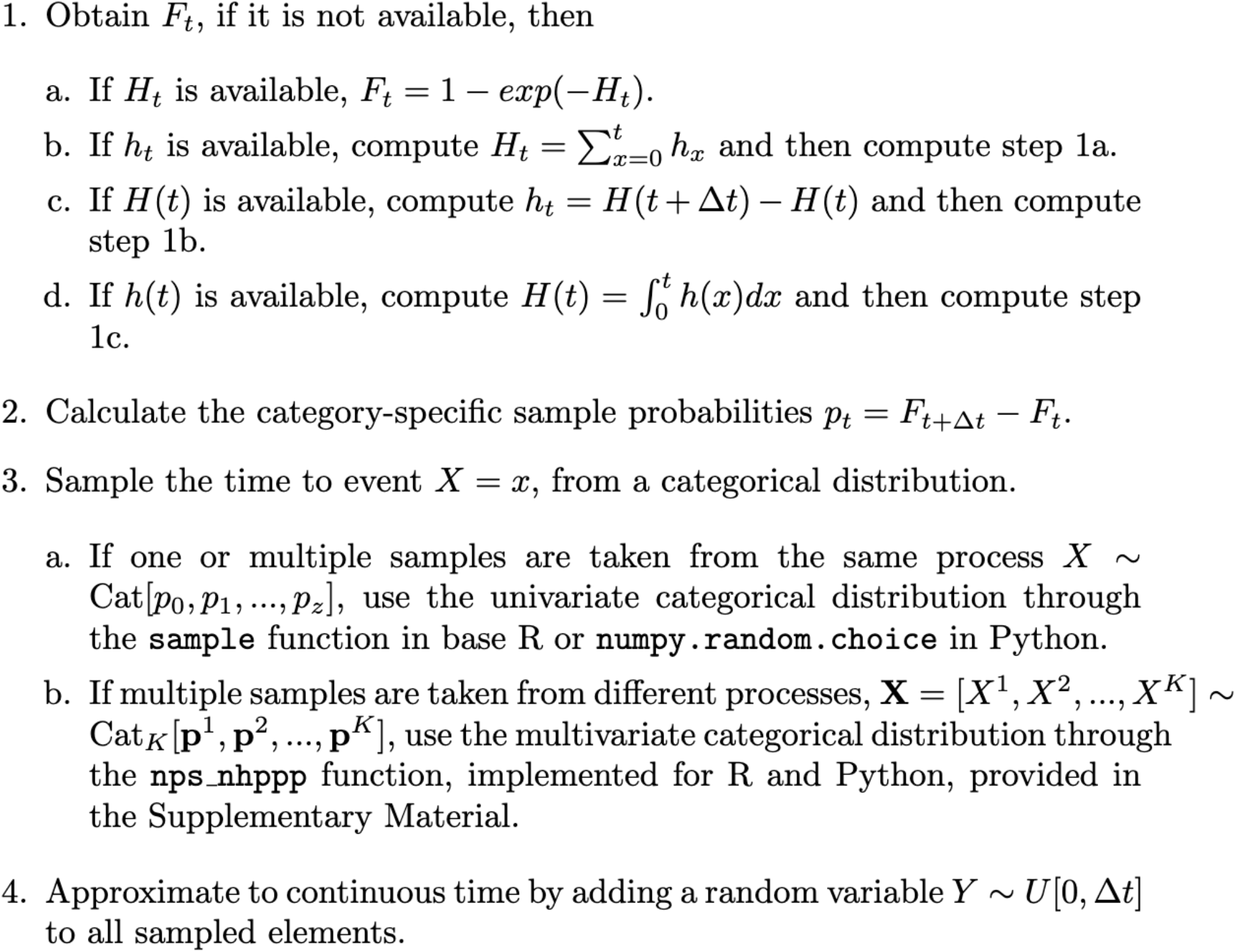
Steps to draw a time-to-event using a nonparametric sampling (NPS) approach. F_t_: cumulative distribution function for a discrete-time random variable; H_t_: discrete-time cumulative hazard function; h_t_: discrete-time hazard function; H(t): continuous time cumulative hazard function; h(t): continuous time hazard function; p_t_: probability mass function; Cat: categorical distribution; and Cat_K_: multivariate categorical distribution for K random variables..

## References

1. Karnon J, Stahl J, Brennan A, Caro JJ, Mar J, Möller J. Modeling using discrete event simulation: A report of the ISPOR-SMDM modeling good research practices task force–4. Medical Decision Making [Internet]. 2012;32(5):701–11. Available from: 10.1177/0272989X12455462

2. Arrospide A, Ibarrondo O, Blasco-Aguado R, Larrañaga I, Alarid-Escudero F, Mar J. Using age-specific rates for parametric survival function estimation in simulation models. Medical Decision Making [Internet]. 2024;0(0):0272989X241232967. Available from: 10.1177/0272989X241232967

3. Cinlar E. Introduction to Stochastic Processes. Englewood Cliffs, NJ: Prentice-Hall; 1975.

4. Trikalinos TA, Sereda Y. Nhppp: Simulating nonhomogeneous poisson point processes in r [Internet]. 2024. Available from: https://arxiv.org/abs/2402.00358

5. Lee ET, Wang JW. Statistical methods for survival data analysis. Hoboken, New Jersey: Wiley; 2013.

6. Klein JP, Moeschberger ML. Survival analysis: Techniques for censored and truncated data. New York, NY: Springer New York; 2003.

7. Elbasha EH, Chhatwal J. Theoretical foundations and practical applications of within-cycle correction methods. Medical Decision Making. 2016;36(1):115–31.

8. Elbasha EH, Chhatwal J. Myths and misconceptions of within-cycle correction: a guide for modelers and decision makers. PharmacoEconomics. 2016;34(1):13–22.

9. Hunink MGGM, Weinstein MC, Wittenberg E, Drummond MF, Pliskin JS, Wong JB, et al. Decision Making in Health and Medicine [Internet]. 2nd ed. Cambridge: Cambridge University Press; 2014. Available from: http://ebooks.cambridge.org/ref/id/CBO9781139506779

10. Hunink MGM, Weinstein MC, Wittenberg E, Drummond MF, Pliskin JS, Wong JB, et al. Decision Making in Health and Medicine: Integrating Evidence and Values. 2nd ed. Cambridge: Cambridge University Press; 2014.

11. Max Planck Institute for Demographic Research (Germany), University of California-Berkeley (USA) and French Institute for Demographic Studies (France). HMD. Human mortality database. https://www.mortality.org/Country/Country?cntr=USA; 2021.

12. Austin PC. Generating survival times to simulate cox proportional hazards models with time-varying covariates. Statistics in Medicine [Internet]. 2012;31(29):3946–58. Available from: 10.1002/sim.5452

13. Julius S. Ngwa DMC Howard J. Cabral, Cupples LA. Generating survival times with time-varying covariates using the lambert w function. Communications in Statistics - Simulation and Computation [Internet]. 2022;51(1):135–53. Available from: 10.1080/03610918.2019.1648822

14. Jeon, J, Meza, R, Krapcho, M, Clarke, L, Byrne, J, Levy, D. “Chapter 5: Actual and counterfactual smoking prevalence rates in the U.S. population via microsimulation.”. Risk Anal 2012; 32 Suppl 1(Suppl 1):S51–68.

15. Willekens F. Multistate analysis of life histories with r [Internet]. Springer International Publishing; 2014. (Use r!). Available from: https://books.google.com.mx/books?id=Cd2CBAAAQBAJ

