## Supplementary material for "A Fast Nonparametric Sampling (NPS) Method for Time-to-Event in Individual-Level Simulation Models"

### Example 1 using R

#### Time to event from parametric hazards

#### Code function

This document presents the code corresponding to the first example presented in the “A Fast Nonparametric Sampling (NPS) Method for Time-to-Event in Individual-Level Simulation Models.” manuscript, all of them using R.

```
# 01 Initial Setup --------------------------------------------------------

# Clean global environment
remove(list = ls())

# Free unused R memory
gc()
```

```
# 02 Define general parameters --------------------------------------------

#´ Number of samples to draw from the life table
n_samp_life_tables <- 1e5

#' Numberof samples, by sex, to draw from the life table
n_samp_by_sex <- 1e5

# Sample size for every sampling iteration
```

[*]School of Epidemiology and Public Health, Faculty of Medicine, University of Ottawa, Ottawa, ON, CA.
[†]School of Epidemiology and Public Health, Faculty of Medicine, University of Ottawa, Ottawa, ON, CA.
[‡]Department of Health Policy, Stanford University School of Medicine, Stanford, CA, USA.
[§]Center for Health Policy, Freeman Spogli Institute, Stanford University, Stanford, CA, USA.

```
n_samples  <- 1e4

# Number of times to repeat the sampling
n_sim_reps <- 1e3

# Number of repetitions in microbenchmark
n_reps_microbench <- 100

# Seed for reproducibility in random number generation
n_seed <- 10242022
```

```
# 03 Define required functions --------------------------------------------

# Calculate expected value using NPS method
calc_ev_nps <- function(n_samples, v_probs){

  # Sample times to event
  v_time_to_event_rates_cat <- sample(size    = n_samples,
                                      x       = 0:150,
                                      prob    = v_probs,
                                      replace = TRUE)

  return(mean(v_time_to_event_rates_cat))
}

# Calculate expected value using NPS approach and continous time approximation
calc_ev_nps_corr <- function(n_samples, v_probs){

  # Sample times to event
  v_time_to_event_rates_cat <- sample(size    = n_samples,
                                      x       = 0:150,
                                      prob    = v_probs,
                                      replace = TRUE)

  # Generate a random number following a unifrom distribution
  v_unif <- runif(n_samples)

  # Add random number
  v_time_to_event_rates_cat_unif <- v_time_to_event_rates_cat + v_unif

  return(mean(v_time_to_event_rates_cat_unif))
}
```

```r
calc_stats_nps_corr <- function(n_reps = 1000,
                                n_samples = 10000, v_time_int = 0:150,
                                v_probs, alpha_level = 0.05,
                                true_ev, true_var){
  # Draw ´n_reps´ samples from the NPS TTT
  m_out <- replicate(n = n_reps,
                     expr = sample_ttt_nps(n_samples = n_samples,
                                           v_time_int = v_time_int,
                                           v_probs = v_probs))

  ## Calculate summary statistics
  v_ev_est  <- colMeans(m_out)
  v_var_est <- matrixStats::colVars(m_out)

  v_se_ev_est  <- sqrt(v_ev_est/n_samples)
  v_se_var_est <- sqrt(v_var_est/n_samples)

  # Expected values
  mean_ev_est <- mean(v_ev_est)
  mean_var_est <- mean(v_var_est)

  # Bias
  bias_ev_est  <- abs(mean(v_ev_est) - true_ev)
  bias_var_est <- abs(mean(v_var_est) - true_var)

  # Monte Carlo Standard Error (MCSE) of Bias
  mcse_bias_ev_est  <- sqrt(
    (sum((v_ev_est - true_ev)^2)/(n_reps - 1))/n_reps)

  mcse_bias_var_est <- sqrt(
    (sum((v_var_est - true_var)^2)/(n_reps - 1))/n_reps)

  # Mean Square Error (MSE)
  mse_ev_est  <- sum((v_ev_est - true_ev)^2)/n_reps
  mse_var_est <- sum((v_var_est - true_var)^2)/n_reps

  # Confidence interval of bias
  z_score <- qnorm(p = 1 - alpha_level/2)
  ci_bias_ev_est <- c(LB = bias_ev_est - z_score*mcse_bias_ev_est,
                      UB = bias_ev_est + z_score*mcse_bias_ev_est)
  ci_bias_var_est <- c(LB = bias_var_est - z_score*mcse_bias_var_est,
                       UB = bias_var_est + z_score*mcse_bias_var_est)
```

```
  # Confidence intervals pf estimates
  chi_score_lb <- qchisq(p = alpha_level/2, df = (n_samples - 1))
  chi_score_ub <- qchisq(p = 1 - alpha_level/2, df = (n_samples - 1))

  m_ci_ev_est <- cbind(LB = v_ev_est - z_score*v_se_ev_est,
                       UB = v_ev_est + z_score*v_se_ev_est)
  # DescTools::MeanCI(v_time_to_event_rates_cat_unif, conf.level = 0.95)
  # confint(lm(v_time_to_event_rates_cat_unif ~ 1))
  m_ci_var_est <- cbind(LB = ((n_samples - 1)*v_var_est)/chi_score_ub,
                        UB = ((n_samples - 1)*v_var_est)/chi_score_lb)
  # DescTools::VarCI(v_time_to_event_rates_cat_unif, conf.level = 0.95)
  # Coverage
  coverage_ev_est  <- mean(true_ev >= m_ci_ev_est[, 1] &
                             true_ev <= m_ci_ev_est[, 2])

  coverage_var_est <- mean(true_var >= m_ci_var_est[, 1] &
                             true_var <= m_ci_var_est[, 2])
  # Output
  return(c(mean_ev_est = mean_ev_est,
           mean_var_est = mean_var_est,
           bias_ev_est = bias_ev_est,
           bias_var_est = bias_var_est,
           coverage_ev_est = coverage_ev_est,
           coverage_var_est = coverage_var_est
  ))
}

sample_ttt_nps <- function(n_samples,
                           v_time_int = 0:150,
                           v_probs){
  v_time_to_event_rates_cat <- sample(size = n_samples, x = v_time_int,
                                      prob = v_probs,
                                      replace = T)
  v_unif <- runif(n_samples)
  v_time_to_event_rates_cat_unif <- v_time_to_event_rates_cat + v_unif
  return(v_time_to_event_rates_cat_unif)
}
```

```
# 04 Calculate expected values from distributions ------------------------

## Exponential distribution ----
```

```r
# Define distribution parameters
par_exp_rate     <- 0.1

# Analytical values
ev_exp           <- 1/par_exp_rate # Analytical expected value
var_exp          <- ev_exp^2       # Analytical variance

# Get instantaneous probability of occurrence
v_prob_exp_rates <- pexp(q = 1:151, rate = par_exp_rate) -
  pexp(q = 0:150, rate = par_exp_rate)

# Simulations using the nps method multiple times
ev_exp_uncorr <- mean(
  replicate(n_sim_reps,
            expr = calc_ev_nps(n_samples = n_samples,
                               v_probs = v_prob_exp_rates)))

# Simulations using the nps method, adding continuous time approximation
ev_exp_corr <- mean(
  replicate(n_sim_reps,
            expr = calc_ev_nps_corr(n_samples = n_samples,
                                    v_probs = v_prob_exp_rates)))

# Round values
ev_exp_corr   <- round(ev_exp_corr, 2)
ev_exp_uncorr <- round(ev_exp_uncorr, 2)

# Measure mean execution time
## Without continuous time correction
l_mbench_exp_uncorr <- microbenchmark::microbenchmark(
  calc_ev_nps(n_samples = n_samples, v_probs = v_prob_exp_rates),
  times = n_reps_microbench,
  unit = "ms")

#* The default output of microbenchmark is in nanoseconds (1/1e-9).
#* The results are converted into miliseconds
t_m_exp_u   <- format(round(mean(l_mbench_exp_uncorr$time/1e6), 2),
                      nsmall = 2)

t_CI_exp_u  <- format(round(quantile(x = l_mbench_exp_uncorr$time/1e6,
                                     probs = c(0.025, 0.975)), 2),
                      nsmall = 2)
```

```r
## With continuous time correction
l_mbench_exp_corr <- microbenchmark::microbenchmark(
  calc_ev_nps_corr(n_samples = n_samples, v_probs = v_prob_exp_rates),
  times = n_reps_microbench,
  unit = "ms")

#* The default output of microbenchmark is in nanoseconds (1/1e-9).
#* The results are converted into miliseconds
t_m_exp_c  <- format(round(mean(l_mbench_exp_corr$time/1e6), 2),
                     nsmall = 2)

t_CI_exp_c <- format(round(quantile(l_mbench_exp_corr$time/1e6,
                                    probs = c(0.025, 0.975)), 2),
                     nsmall = 2)


# Using general function
v_sim_nps_out_exp <- calc_stats_nps_corr(n_reps      = n_sim_reps,
                                         n_samples   = n_samples,
                                         v_time_int  = 0:150,
                                         v_probs     = v_prob_exp_rates,
                                         alpha_level = 0.05,
                                         true_ev     = ev_exp,
                                         true_var    = var_exp)

## Gamma distribution ----

# Define distribution parameters
par_gamma_shape <- 4
par_gamma_rate <- 0.1

# Analytical values
ev_gamma  <- par_gamma_shape/par_gamma_rate     # Analytical expected value
var_gamma <- par_gamma_shape/(par_gamma_rate^2) # Analytical variance

# Get instantaneous probability of ocurrence
v_prob_gamma_rates <-
  pgamma(q = 1:151, shape = par_gamma_shape, rate = par_gamma_rate) -
  pgamma(q = 0:150, shape = par_gamma_shape, rate = par_gamma_rate)

# Simulations using the nps method multiple times
ev_gamma_uncorr <- mean(
```

```r
  replicate(n_sim_reps,
            expr = calc_ev_nps(n_samples = n_samples,
                               v_probs = v_prob_gamma_rates)))

# Simulations using the nps method, adding continuous time approximation
ev_gamma_corr <- mean(
  replicate(n_sim_reps,
            expr = calc_ev_nps_corr(n_samples = n_samples,
                                    v_probs = v_prob_gamma_rates)))

# Round values
ev_gamma_corr   <- round(ev_gamma_corr, 2)
ev_gamma_uncorr <- round(ev_gamma_uncorr, 2)

# Measure mean execution time
## Without continuous time correction
l_mbench_gamma_uncorr <- microbenchmark::microbenchmark(
  calc_ev_nps(n_samples = n_samples, v_probs = v_prob_gamma_rates),
  times = n_reps_microbench,
  unit = "ms")

#* The default output of microbenchmark is in nanoseconds (1/1e-9).
#* The results are converted into miliseconds
t_m_gamma_u  <- format(round(mean(l_mbench_gamma_uncorr$time/1e6), 2),
                       nsmall = 2)

t_CI_gamma_u <- format(round(quantile(l_mbench_gamma_uncorr$time/1e6,
                                      probs = c(0.025, 0.975)), 2),
                       nsmall = 2)

## With continuous time correction
l_mbench_gamma_corr <- microbenchmark::microbenchmark(
  calc_ev_nps_corr(n_samples = n_samples, v_probs = v_prob_gamma_rates),
  times = n_reps_microbench,
  unit = "ms")

#* The default output of microbenchmark is in nanoseconds (1/1e-9).
#* The results are converted into miliseconds
t_m_gamma_c  <- format(round(mean(l_mbench_gamma_corr$time/1e6), 2),
                       nsmall = 2)

t_CI_gamma_c <- format(round(quantile(l_mbench_gamma_corr$time/1e6,
```

```
                                    probs = c(0.025, 0.975)), 2),
                      nsmall = 2)

# Using general function
v_sim_nps_out_gamma <- calc_stats_nps_corr(n_reps = n_sim_reps,
                                           n_samples = n_samples,
                                           v_time_int = 0:150,
                                           v_probs = v_prob_gamma_rates,
                                           alpha_level = 0.05,
                                           true_ev = ev_gamma,
                                           true_var = var_gamma)


## Log-normal distribution ----

# Define distribution parameters
par_lnorm_meanlog <- 3.5
par_lnorm_sdlog   <- 0.15

# Analytical values
## Analytical expected value
ev_lnorm  <- exp(par_lnorm_meanlog + ((par_lnorm_sdlog^2)/2))

## Analytical variance
var_lnorm <- exp(2*par_lnorm_meanlog + (par_lnorm_sdlog^2))*(
  exp(par_lnorm_sdlog^2) - 1)

# Round values
ev_lnorm <- round(ev_lnorm, 2)

# Get instantaneous probability of ocurrence
v_prob_lnorm_rates <-
  plnorm(q = 1:151, meanlog = par_lnorm_meanlog, sdlog = par_lnorm_sdlog) -
  plnorm(q = 0:150, meanlog = par_lnorm_meanlog, sdlog = par_lnorm_sdlog)

# Simulations using the nps method multiple times
ev_lnorm_uncorr <- mean(
  replicate(n_sim_reps,
            expr = calc_ev_nps(n_samples = n_samples,
                               v_probs = v_prob_lnorm_rates)))

# Simulations using the nps method, adding continuous time approximation
```

```
ev_lnorm_corr <- mean(
  replicate(n_sim_reps,
            expr = calc_ev_nps_corr(n_samples = n_samples,
                                    v_probs = v_prob_lnorm_rates)))


# Round values
ev_lnorm_corr   <- round(ev_lnorm_corr, 2)
ev_lnorm_uncorr <- round(ev_lnorm_uncorr, 2)

# Measure mean execution time
## Without continuous time correction
l_mbench_lnorm_uncorr <- microbenchmark::microbenchmark(
  calc_ev_nps(n_samples = n_samples, v_probs = v_prob_lnorm_rates),
  times = n_reps_microbench,
  unit = "ms")

#* The default output of microbenchmark is in nanoseconds (1/1e-9).
#* The results are converted into miliseconds
t_m_lnorm_u  <- format(round(mean(l_mbench_lnorm_uncorr$time/1e6), 2),
                       nsmall = 2)

t_CI_lnorm_u <- format(round(quantile(l_mbench_lnorm_uncorr$time/1e6,
                                      probs = c(0.025, 0.975)), 2),
                       nsmall = 2)

## With continuous time correction
l_mbench_lnorm_corr <- microbenchmark::microbenchmark(
  calc_ev_nps_corr(n_samples = n_samples, v_probs = v_prob_lnorm_rates),
  times = n_reps_microbench,
  unit = "ms")

#* The default output of microbenchmark is in nanoseconds (1/1e-9).
#* The results are converted into miliseconds
t_m_lnorm_c  <- format(round(mean(l_mbench_lnorm_corr$time/1e6), 2),
                       nsmall = 2)

t_CI_lnorm_c <- format(round(quantile(l_mbench_lnorm_corr$time/1e6,
                                      probs = c(0.025, 0.975)), 2),
                       nsmall = 2)

### Using general function
```

```r
v_sim_nps_out_lnorm <- calc_stats_nps_corr(n_reps      = n_sim_reps,
                                           n_samples   = n_samples,
                                           v_time_int  = 0:150,
                                           v_probs     = v_prob_lnorm_rates,
                                           alpha_level = 0.05,
                                           true_ev     = ev_lnorm,
                                           true_var    = var_lnorm)

# Remove seed
set.seed(NULL)
```

### Example 2 using R

#### Drawing age to death from homogeneous cohort

David Garibay, M.P.P.[*] Hawre Jalal, MD, Ph.D.[†]
Fernando Alarid-Escudero, Ph.D.[‡§]

#### Code function

This document presents the code corresponding to the second example presented in the "A Fast Nonparametric Sampling (NPS) Method for Time-to-Event in Individual-Level Simulation Models." manuscript, all of them using R.

```
# 01 Initial Setup ---------------------------------------------------------

# Clean global environment
remove(list = ls())

# Free unused R memory
gc()

# Load libraries
library(dplyr)
library(ggplot2)
library(tidyr)
library(tibble)
library(microbenchmark)
```

[*]School of Epidemiology and Public Health, Faculty of Medicine, University of Ottawa, Ottawa, ON, CA.
[†]School of Epidemiology and Public Health, Faculty of Medicine, University of Ottawa, Ottawa, ON, CA.
[‡]Department of Health Policy, Stanford University School of Medicine, Stanford, CA, USA.
[§]Center for Health Policy, Freeman Spogli Institute, Stanford University, Stanford, CA, USA.

```r
# 02 Define general parameters --------------------------------------------

# Number of samples to draw from the life table
n_samp_life_tables <- 1e5

# Number of iterations for microbenchmarking
n_samp_iter_life_tables <- 100

# Seed for reproducibility in random number generation
n_seed <- 10242022

# To print a specific number of digits in tibbles
options(pillar.sigfig = 4)
```

```r
# 03 Load base data -------------------------------------------------------

#* Yearly USA data, from 2000 to 2019,
#* Mortality rate for males, females and total
#* Obtained from The Human Mortality Database:
#* https://www.mortality.org/cgi-bin/hmd/country.php?cntr=USA&level=1
load("../data/all_cause_mortality.rda")
```

```r
# 04 Filter data ----------------------------------------------------------

# For homogeneous population example
df_all_cause_mortality_filt <- all_cause_mortality %>%
  as_tibble() %>%
  filter(Year == 2015)
```

```r
# 05 Data wrangling -------------------------------------------------------

#* Following Lee & Wang (2013)-Statistical methods for survival data analysis
#* 4th ed - chapter 2: Functions of survival time
df_lifetable <- df_all_cause_mortality_filt %>%
  dplyr::arrange(Sex, Year, Age) %>%
  dplyr::group_by(Sex) %>%
  dplyr::mutate(
    H_t = cumsum(Rate),        # H(t) - Cumulative hazard
    S_t = exp(-H_t),           # S(t) - Cumulative survival
    F_t = 1 - exp(-H_t),       # F(t) - Cumulative probability: 1 - S(t)
    p_t = c(F_t[1], diff(F_t)) # f(t) - Instantaneous probability
  ) %>%
```

```
  ungroup()


# Calculate life expectancy from lifetables data
df_le_lifetable <- df_lifetable %>%
  group_by(Sex) %>%
  summarise(le = sum(S_t))


# Obtain life expectancy from lifetables
le_lifetable_homog <- df_le_lifetable[df_le_lifetable$Sex == "Total", ]$le
```

```
# Set seed for reproducibility in random number generation
set.seed(n_seed)

# 06 Calculate life expectancy using nps method ---------------------------

# Filter to have homogeneous population
df_lifetable_homog <- df_lifetable %>%
  filter(Sex == "Total")


#* Sample ages to death from a categorical sampling
v_cat_life_table_homog <- sample(x       = df_lifetable_homog$Age,
                                 size    = n_samp_life_tables,
                                 prob    = df_lifetable_homog$p_t,
                                 replace = TRUE)

#* Create vector of drawings following a uniform distribution
v_unif_life_table_homog <- runif(n = n_samp_life_tables, min = 0, max = 1)

#* Add this vector to the categorical sampling outputs
v_cat_life_table_corr_homog <- (v_cat_life_table_homog +
                                  v_unif_life_table_homog)

#* Life expectancy without continuous time correction
le_homog_uncorr <- mean(v_cat_life_table_homog)

#* Life expectancy with correction
le_homog_corr <- mean(v_cat_life_table_corr_homog)

# Measure mean execution time
```

```r
## Without continuous time correction
l_mbench_homog_uncorr <- microbenchmark::microbenchmark(
  sample(x       = df_lifetable_homog$Age,
         size    = n_samp_life_tables,
         prob    = df_lifetable_homog$p_t,
         replace = TRUE),
  times = n_samp_iter_life_tables,
  unit = "ms")

## With continuous time correction
l_mbench_homog_corr <- microbenchmark::microbenchmark(
  sample(x       = df_lifetable_homog$Age,
         size    = n_samp_life_tables,
         prob    = df_lifetable_homog$p_t,
         replace = TRUE) + runif(n = n_samp_life_tables, min = 0, max = 1),
  times = n_samp_iter_life_tables,
  unit = "ms")


# Remove seed
set.seed(NULL)
```

### Example 3 using R

#### Drawing age to death from a heterogeneous cohort

David Garibay, M.P.P.[*] Hawre Jalal, MD, Ph.D.[†]
Fernando Alarid-Escudero, Ph.D.[‡§]

#### Code function

This document presents the code corresponding to the third example presented in the "A Fast Nonparametric Sampling (NPS) Method for Time-to-Event in Individual-Level Simulation Models." manuscript, all of them using R.

```
# 01 Initial Setup --------------------------------------------------------

## 01.01 Clean environment ------------------------------------------------
remove(list = ls())

#* Refresh environment memory
gc()

## 01.02 Load libraries ----------------------------------------------------
library(dplyr)
library(ggplot2)
library(tidyr)
library(tibble)
library(microbenchmark)

# Load function to implement multivariate categorical sampling
source(file = "../R/nps_nhppp.R")
```

[*]School of Epidemiology and Public Health, Faculty of Medicine, University of Ottawa, Ottawa, ON, CA.
[†]School of Epidemiology and Public Health, Faculty of Medicine, University of Ottawa, Ottawa, ON, CA.
[‡]Department of Health Policy, Stanford University School of Medicine, Stanford, CA, USA.
[§]Center for Health Policy, Freeman Spogli Institute, Stanford University, Stanford, CA, USA.

```
# 02 Define general parameters -------------------------------------------

# Number of samples to draw from the life table
n_samp_life_tables <- 1e5

# Number of samples, by sex, to draw from the life table
n_samp_by_sex <- 1e5

# Number of iterations for microbenchmarking
n_samp_iter_life_tables <- 100

# Seed for reproducibility in random number generation
n_seed <- 10242022

# To print a specific number of digits in tibbles
options(pillar.sigfig = 4)
```

```
# 03 Load base data -------------------------------------------------------

#* Yearly USA data, from 2000 to 2019,
#* Mortality rate for males, females and total
#* Obtained from The Human Mortality Database:
#* https://www.mortality.org/cgi-bin/hmd/country.php?cntr=USA&level=1
load("../data/all_cause_mortality.rda")
```

```
# 04 Filter data ----------------------------------------------------------

# For homogeneous population example
df_all_cause_mortality_filt <- all_cause_mortality %>%
  as_tibble() %>%
  filter(Year == 2015)
```

```
# 05 Data wrangling -------------------------------------------------------

#* Following Lee & Wang (2013)-Statistical methods for survival data analysis
#* 4th ed - chapter 2: Functions of survival time
df_lifetable <- df_all_cause_mortality_filt %>%
  dplyr::arrange(Sex, Year, Age) %>%
  dplyr::group_by(Sex) %>%
  dplyr::mutate(
    H_t = cumsum(Rate),        # H(t) - Cumulative hazard
    S_t = exp(-H_t),           # S(t) - Cumulative survival
```

```
    F_t = 1 - exp(-H_t),        # F(t) - Cumulative probability: 1 - S(t)
    p_t = c(F_t[1], diff(F_t)) # f(t) - Instantaneous probability
  ) %>%
  ungroup()


# Calculate life expectancy from lifetables data
df_le_lifetable <- df_lifetable %>%
  group_by(Sex) %>%
  summarise(le = sum(S_t))


# Obtain life expectancy by sex from life tables
le_lifetable_fem  <- df_le_lifetable[df_le_lifetable$Sex == "Female",]$le
le_lifetable_male <- df_le_lifetable[df_le_lifetable$Sex == "Male",]$le
```

```
# Seed for reproducibility in random number generation
set.seed(n_seed)

# 06 Calculate life expectancy using nps method ---------------------------

v_samp_sex <- c("Male", "Female")

# Filter to have heterogeneous population
df_lifetable_heterog <- df_lifetable %>%
  filter(Sex != "Total") %>%
  select(Year, Sex, Age, p_t) %>%
  # Normalize probabilities by sex
  group_by(Sex) %>%
  # Normalize instantaneous probabilities by sex
  mutate(p_t = p_t / sum(p_t)) %>%
  ungroup()

# Generate synthetic cohort with 50% males and 50% females
df_samp_raw <- tibble::tibble(
  Year = 2015,
  Sex = c(rep(x    = v_samp_sex,
              each = n_samp_by_sex)))

# Convert lifetable data from long to wide
df_lifetable_probs_wide <- tidyr::pivot_wider(
  data = df_lifetable_heterog,
```

```
  names_from = Age,
  values_from = p_t,
  names_prefix = "Age_")

# Generate dataset for Multivariate categorical sampling
df_samp_probs <- df_samp_raw %>%
  left_join(y = df_lifetable_probs_wide,
            by = join_by(Year, Sex))

#* Extract probability matrix from `df_samp_probs`
m_probs <- df_samp_probs %>%
  select(-Year, -Sex) %>%
  as.matrix()

#* Implement Multivariate NPS
##* Without continuous time approximation
v_cat_life_table_heterog <- nps_nhppp(m_probs = m_probs,
                                      correction = "none")

##* With continuous time approximation
v_cat_life_table_heterog_corr <- nps_nhppp(m_probs = m_probs,
                                           correction = "uniform")

# Create dataset with the age to death samples
df_heterog_samp <- df_samp_raw %>%
  mutate(age_death      = v_cat_life_table_heterog,
         age_death_corr = v_cat_life_table_heterog_corr,
         .after = Sex)

#* Obtain life expectancies by sex
df_le_nps_heterog <- df_heterog_samp %>%
  group_by(Sex) %>%
  summarise(le = mean(age_death),
            le_corr = mean(age_death_corr)) %>%
  ungroup()


# Extract values
le_nps_fem_uncorr  <- filter(df_le_nps_heterog, Sex == "Female")$le
le_nps_male_uncorr <- filter(df_le_nps_heterog, Sex == "Male")$le

le_nps_fem_corr   <- filter(df_le_nps_heterog, Sex == "Female")$le_corr
```

```r
le_nps_male_corr   <- filter(df_le_nps_heterog, Sex == "Male")$le_corr

# Measure mean execution time
## Without continuous time correction
l_mbench_heterog_uncorr <- microbenchmark::microbenchmark(
  nps_nhppp(m_probs = m_probs, correction = "none"),
  times = n_samp_iter_life_tables,
  unit = "ms")

## With continuous time correction
l_mbench_heterog_corr <- microbenchmark::microbenchmark(
  nps_nhppp(m_probs = m_probs, correction = "uniform"),
  times = n_samp_iter_life_tables,
  unit = "ms")

# Remove seed
set.seed(NULL)
```

### Example 4 using R

**Drawing time to event from hazards with time-dependent covariates**

David Garibay, M.P.P.[*] Hawre Jalal, MD, Ph.D.[†]
Fernando Alarid-Escudero, Ph.D.[‡§]

#### Code function

This document presents the code corresponding to the fourth example presented in the "A Fast Nonparametric Sampling (NPS) Method for Time-to-Event in Individual-Level Simulation Models." manuscript, all of them using R.

```
# 01 Initial Setup ---------------------------------------------------------

## 01.01 Clean environment ------------------------------------------------
remove(list = ls())

#* Refresh environment memory
gc()

# 01.02 Load libraries ----------------------------------------------------
library(dplyr)
library(ggplot2)
library(tidyr)
library(tibble)
library(data.table)
library(flexsurv)
library(LambertW)
library(reshape2)
library(microbenchmark)
```

[*]School of Epidemiology and Public Health, Faculty of Medicine, University of Ottawa, Ottawa, ON, CA.
[†]School of Epidemiology and Public Health, Faculty of Medicine, University of Ottawa, Ottawa, ON, CA.
[‡]Department of Health Policy, Stanford University School of Medicine, Stanford, CA, USA.
[§]Center for Health Policy, Freeman Spogli Institute, Stanford University, Stanford, CA, USA.

```
# Load function to implement multivariate categorical sampling
source(file = "../R/nps_nhppp.R")
```

```
# 02 Define general parameters -------------------------------------------

# Parameters for time-varying covariates
alpha_0 <- 0
alpha_1 <- 1
# When beta <- 0 the time-varying covariate is deactivated
beta    <- log(1.02)

# Define a time range
time_var_cov <- seq(0, 100)

# Sample size
n_samp <- 1e6

#* Number of iterations for microbenchmarking in time-dependent covariates
#* examples
n_iter_time_var_cov <- 100

# Seed for reproducibility in random number generation
n_seed <- 10242022
```

```
# 03 Define required functions -------------------------------------------

## Function to apply the time-varying covariate to a baseline hazard
compute_time_varying_hazard_linear_2 <- function(hazard0,
                                                 alpha_0,
                                                 alpha_1,
                                                 beta,
                                                 time_var_cov){

  # With this function we take the diagonal values of the matrix
  hazard <- hazard0*exp(beta*(alpha_0 + alpha_1*time_var_cov))

  # #* With this specification we obtain the full matrix and then we need to
  # #* make a subset considering a certain set of time-varying covariates
  # #* - DEACTIVATED
  # hazard <- hazard0 %*% exp(beta*(alpha_0 + alpha_1*time_var_cov))
```

```r
  return(hazard)
}


#* Function to get time to events from exponential baseliine hazard
#* using analytic formula following Austin's 2012 equations:
#* - Austin, P. C. (2012).
#*   Generating survival times to simulate Cox proportional hazards models
#*   with time-varying covariates. Statistics in Medicine, 31(29), 3946-3958.
#*   https://doi.org/10.1002/sim.5452
inv_exp_time_ <- function(n_samp, rate, alpha_0, alpha_1, beta) {

  v_unif <- runif(n = n_samp)

  exp_time <- (1/(beta*alpha_1))*log(1 +
                                       ((-alpha_1*beta*log(v_unif))/
                                          (rate*exp(alpha_0*beta)))
  )

  return(exp_time)
}

#* Function to get time to events from Gompertz baseliine hazard
#* using analytic formula following Austin's 2012 equations
inv_gomp_time_ <- function(n_samp,
                           shape,
                           rate,
                           alpha_0,
                           alpha_1,
                           beta) {

  v_unif <- runif(n = n_samp)

  gomp_time <- (1/((beta*alpha_1) + shape))*
    log(1 + (((beta*alpha_1) + shape)*(-log(v_unif)))/
          (rate*exp(alpha_0*beta)))

  return(gomp_time)
}

#* Function to get time to events from Gompertz baseliine hazard
#* using analytic formula following Ngwa, et al.'s equations:
```

```r
#* - Ngwa, J. S., Cabral, H. J., Cheng, D. M., Gagnon, D. R., LaValley,
#*   M. P., & Cupples, L. A. (2022). Generating survival times with
#*   time-varying covariates using the Lambert W Function. Communications in
#*   Statistics: Simulation and Computation, 51(1), 135-153.
#    https://doi.org/10.1080/03610918.2019.1648822
inv_weibull_time <- function(n_samp,
                             shape,
                             scale,
                             alpha_0,
                             alpha_1,
                             beta) {

  v_unif <- runif(n = n_samp)

  weibull_time <- (1/(beta*alpha_1*(1/shape)))*LambertW::W(
    beta*(alpha_1*(1/shape))*(
      -log(v_unif)/scale*exp(beta*alpha_0)
    )^(1/shape)
  )

  return(weibull_time)
}
```

```r
# 04 Draw time to events ---------------------------------------------------

# Add seed for reproducibility
set.seed(n_seed)

## 04.01 Exponential baseline hazard ---------------------------------------

# Define general parameters for the Exponential baseline hazard
rate    <- 0.1

# Obtain the exponential baseline hazard
v_exp_hazard0 <- matrix(data = flexsurv::hexp(x = 0:100, rate = rate),
                        ncol = 1)

#* Compute the hazard after adding a proportional hazards approach using a
#* time-varying covariate
hazard <- compute_time_varying_hazard_linear_2(
  hazard0      = v_exp_hazard0,
  alpha_0      = alpha_0,
```

```
  alpha_1      = alpha_1,
  beta         = beta,
  time_var_cov = time_var_cov)


df_hazard_long <- reshape2::melt(data       = hazard,
                                 varnames   = c("Time", "Covariate"),
                                 value.name = "h(t)")

dt_hazard_long <- data.table::as.data.table(df_hazard_long)

#* Transform hazards `h(t)` to instantaneous probabilities `f(t)`
#* # H(t) - Cumulative hazard
dt_hazard_long[, H := cumsum(`h(t)`)]
# F(t) - Cumulative probability
dt_hazard_long[, `F` := 1 - exp(-H)]
# f(t) - Instantaneous probability
dt_hazard_long[,  f := c(`F`[1], diff(`F`))]

#* Sample times to event considering the previously defined instantaneous
#* probabilities
v_time_to_event_random_path <- sample(x    = time_var_cov,
                                      size = n_samp,
                                      prob = dt_hazard_long$f,
                                      replace = TRUE)

# Add continous time approximation
v_time_to_event_random_path <-  v_time_to_event_random_path +
  runif(n = length(v_time_to_event_random_path))


# Obtain times to event following analytical formula
v_exp_time <- inv_exp_time_(n_samp  = n_samp,
                            rate    = rate,
                            alpha_0 = alpha_0,
                            alpha_1 = alpha_1,
                            beta    = beta)

# Compare mean time to event of
## Analytical formula sample
ev_exp_time_af <- mean(v_exp_time)
## NPS method
```

```r
ev_exp_time_nps <- mean(v_time_to_event_random_path)

# Measure mean execution time
l_mbench_tvar_exp <- microbenchmark::microbenchmark(
  sample(x = 0:100,
         size = 1e6,
         prob = dt_hazard_long$f,
         replace = TRUE),
  times = n_iter_time_var_cov,
  unit = "ms")

## 04.02 Gompertz baseline hazard ------------------------------------------

# Define general parameters for the Weibull baseline hazard
shape <- 0.1
rate <- 0.001

# Obtain the Gompertz baseline hazard
v_gomp_hazard0 <- flexsurv::hgompertz(x = 0:100, shape = shape, rate = rate)

#* Compute the hazard after adding a proportional hazards approach using a
#* time-varying covariate
gomp_hazard <- compute_time_varying_hazard_linear_2(
  hazard0      = v_gomp_hazard0,
  alpha_0      = alpha_0,
  alpha_1      = alpha_1,
  beta         = beta,
  time_var_cov = time_var_cov)

df_gomp_hazard_long <- reshape2::melt(data       = gomp_hazard,
                                      varnames   = c("Time", "Covariate"),
                                      value.name = "h(t)")

dt_gomp_hazard_long <- as.data.table(df_gomp_hazard_long)

#* Transform hazards `h(t)` to instantaneous probabilities `f(t)`
dt_gomp_hazard_long[, H := cumsum(`h(t)`)]
dt_gomp_hazard_long[, `F` := 1 - exp(-H)]
dt_gomp_hazard_long[, f := c(`F`[1], diff(`F`))]

# Sample times to event considering the previously defined instantaneous
#probabilities
```

```
v_time_to_event_gompertz <- sample(x = time_var_cov,
                                   size = n_samp,
                                   prob = dt_gomp_hazard_long$f,
                                   replace = TRUE)

# Add continous time approximation
v_time_to_event_gompertz <-  v_time_to_event_gompertz +
  runif(n = length(v_time_to_event_gompertz))

# Obtain times to event following analytical formula
v_gomp_time <- inv_gomp_time_(n_samp  = n_samp,
                              shape   = shape,
                              rate    = rate,
                              alpha_0 = alpha_0,
                              alpha_1 = alpha_1,
                              beta    = beta)


# Compare expected time to event of
## Analytical formula sample
ev_gomp_time_af <- mean(v_gomp_time)
## Proposed approach
ev_gomp_time_nps <- mean(v_time_to_event_gompertz)

# Measure mean execution time
l_mbench_tvar_gomp <- microbenchmark(
  sample(x = 0:100,
         size = 1e6,
         prob = dt_gomp_hazard_long$f,
         replace = T) +
    runif(n = length(v_time_to_event_gompertz)),
  times = n_iter_time_var_cov,
  unit = "ms")


## 04.03 Weibull baseline hazard -------------------------------------------

# Define general parameters for the Weibull baseline hazard
n_shape_weib = 2
n_scale_weib = 0.01

# Obtain the Weibull (proportional hazards) baseline hazard
```

```
v_weibull_hazard0 <- matrix(flexsurv::hweibullPH(x     = time_var_cov,
                                                 shape = n_shape_weib,
                                                 scale = n_scale_weib))

#* Compute the hazard after adding a proportional hazards approach using a
#* time-varying covariate
weibull_hazard <- compute_time_varying_hazard_linear_2(
  hazard0      = v_weibull_hazard0,
  alpha_0      = alpha_0,
  alpha_1      = alpha_1,
  beta         = beta,
  time_var_cov = time_var_cov)

df_weibull_hazard_long <- reshape2::melt(data       = weibull_hazard,
                                         varnames   = c("Time", "Covariate"),
                                         value.name = "h(t)")

dt_weibull_hazard_long <- as.data.table(df_weibull_hazard_long)

# Sample time to events for random path
dt_weibull_hazard_long[, H := cumsum(`h(t)`)]
dt_weibull_hazard_long[, `F` := 1 - exp(-H)]
dt_weibull_hazard_long[, f := c(`F`[1], diff(`F`))]

# Sample times to event considering the previously defined instantaneous
# probabilities
v_time_to_event_weibull <- sample(x       = time_var_cov,
                                  size    = n_samp,
                                  prob    = dt_weibull_hazard_long$f,
                                  replace = TRUE)

# Add continous time approximation
v_time_to_event_weibull <-  v_time_to_event_weibull +
  runif(n = length(v_time_to_event_weibull))

# Obtain times to event following analytical formula
v_weibull_time <- inv_weibull_time(n_samp  = n_samp,
                                   shape   = n_shape_weib,
                                   scale   = n_scale_weib,
                                   alpha_0 = alpha_0,
                                   alpha_1 = alpha_1,
                                   beta    = beta)
```

```
# Compare expected time to event of
## Analytical formula sample
ev_weibull_time_af <- mean(v_weibull_time)
## Proposed approach
ev_weibull_time_nps <- mean(v_time_to_event_weibull)

# Measure mean execution time
l_mbench_tvar_weib <- microbenchmark::microbenchmark(
  sample(x = 0:100,
         size = 1e6,
         prob = dt_weibull_hazard_long$f,
         replace = T) + runif(n = length(v_time_to_event_weibull)),
  times = n_iter_time_var_cov,
  unit = "ms")

# Remove seed
set.seed(NULL)
```

### Example 5 using R

**Drawing time to event from hazards with time-dependent covariates following random paths**

David Garibay, M.P.P.[*] Hawre Jalal, MD, Ph.D.[†]
Fernando Alarid-Escudero, Ph.D.[‡§]

#### Code function

This document presents the code corresponding to the fifth example presented in the “A Fast Nonparametric Sampling (NPS) Method for Time-to-Event in Individual-Level Simulation Models.” manuscript, all of them using R.

```
# 01 Initial Setup --------------------------------------------------------

## 01.01 Clean environment ------------------------------------------------
remove(list = ls())

#* Refresh environment memory
gc()

# 01.02 Load libraries ----------------------------------------------------
library(dplyr)
library(ggplot2)
library(tidyr)
library(tibble)
library(data.table)
library(flexsurv)
library(LambertW)
library(reshape2)
```

[*]School of Epidemiology and Public Health, Faculty of Medicine, University of Ottawa, Ottawa, ON, CA.
[†]School of Epidemiology and Public Health, Faculty of Medicine, University of Ottawa, Ottawa, ON, CA.
[‡]Department of Health Policy, Stanford University School of Medicine, Stanford, CA, USA.
[§]Center for Health Policy, Freeman Spogli Institute, Stanford University, Stanford, CA, USA.

```r
library(microbenchmark)

# Load function to implement multivariate categorical sampling
source(file = "../R/nps_nhppp.R")
```

```r
# 02 Define general parameters ---------------------------------------------

# Parameters for time-varying covariates
alpha_0 <- 0
alpha_1 <- 1
# When beta <- 0 the time-varying covariate is deactivated
beta    <- log(1.005)


# Define parameters for the Weibull baseline hazard
n_weib_shape <- 1.3
n_weib_scale <- 30.1

n_ind    <-  1000 # Number of simulated individuals
n_cycles <-  100  # Number of cycles
ourDrift <-  0.5

#* Number of iterations for microbenchmarking in time-dependent covariates
#* examples
n_iter_time_var_cov <- 100

# Seed for reproducibility in random number generation
n_seed <- 10242022
```

```r
# 03 Define required functions ---------------------------------------------

# Define random path function
create_time_varying_covariate <- function(n_ind    = 100,
                                          n_cycles = 100,
                                          ourDrift = 0.005){

  m_random_paths <- matrix(nrow = n_ind, ncol = n_cycles)
  m_random_paths[, 1] <- 1

  for (cycle in 2:n_cycles) {

    v_next_step = rnorm(n = n_ind, mean = 0, sd = ourDrift)
```

```
    m_random_paths[, cycle] <- round(pmax(m_random_paths[, cycle - 1] +
                                          v_next_step, 0))
  }

  dtb_paths_individuals <- as.data.table(
    reshape2::melt(data       = m_random_paths,
                   varnames   = c("id", "Time"),
                   value.name = "Covariate"))

  setorder(dtb_paths_individuals, id, Time)

  return(dtb_paths_individuals)
}

## Function to apply the time-varying covariate to a baseline hazard
compute_time_varying_hazard_linear_3 <- function(hazard0,
                                                 alpha_0,
                                                 alpha_1,
                                                 beta,
                                                 time_var_cov){

  # This specification gets the full matrix of values as output
  hazard <- hazard0 %*% exp(beta*(alpha_0 + alpha_1*time_var_cov))

  return(hazard)
}
```

```
# 04 Draw time to events ----------------------------------------------------

# Set seed for reproducibility
set.seed(n_seed)

# Sample from Weibull baseline hazard (h_0)
hazard0 <- matrix(flexsurv::hweibull(x     = 1:n_cycles,
                                     shape = n_weib_shape,
                                     scale = n_weib_scale),
                  ncol = 1)


# Compute values of the hazard based on a range of covariate values
weibull_hazard <- compute_time_varying_hazard_linear_3(
  hazard0      = hazard0,
```

```
  alpha_0      = alpha_0,
  alpha_1      = alpha_1,
  beta         = beta,
  time_var_cov = seq(0:n_cycles))

# Convert to long format
df_weibull_hazard_long <- reshape2::melt(data       = weibull_hazard,
                                         varnames   = c("Time", "Covariate"),
                                         value.name = "h(t)")

dt_weibull_hazard_long <- as.data.table(df_weibull_hazard_long)

# Correct covariate id
dt_weibull_hazard_long[, Covariate := Covariate - 1]

#* Set key for efficient binary search
#* Check `vignette("datatable-keys-fast-subset") `
setkey(dt_weibull_hazard_long, Time, Covariate)

# Create time varying covariate, y_i(t)
dtb_paths_individuals <- create_time_varying_covariate(n_ind    = n_ind,
                                                       n_cycles = n_cycles,
                                                       ourDrift = ourDrift)




# Obtain time-dependent hazards from indvidual-specific random paths
dtb_paths_individuals[, `h(t)` := dt_weibull_hazard_long[
  .(dtb_paths_individuals$Time, dtb_paths_individuals$Covariate),  `h(t)`]]


# Steps to get time-specific probability of event occurrence
# H(t) - Cumulative hazard
dtb_paths_individuals[, H := cumsum(`h(t)`),  by = id]
# F(t) - Cumulative probability
dtb_paths_individuals[, `F` := 1 - exp(-H)]
# f(t) - Instantaneous probability
dtb_paths_individuals[, f := c(`F`[1], diff(`F`)), by = id]

# Generate data set to sample time to event
dt_paths_individuals_wide <- data.table::dcast(data = dtb_paths_individuals,
```

```
                                              value.var = "f",
                                              formula = id ~ Time)

# Generate last cycle to sum probability up to 1
dt_paths_individuals_wide[
  , `101` := 1 - dtb_paths_individuals[Time == 100, `F`]]

# Sample time to event for all individuals
out_nps <- nps_nhppp(
  m_probs = as.matrix(dt_paths_individuals_wide[, `1`:`101`]),
  v_categories = seq(0, 100),
  correction = "none")

# Measure mean execution time
l_mbench_random_path <- microbenchmark::microbenchmark(
  nps_nhppp(m_probs    = as.matrix(dt_paths_individuals_wide[, 2:102]),
            correction = "uniform"),
  times = n_iter_time_var_cov,
  unit = "ms")

# Remove seed
set.seed(NULL)
```

### Definition of the NPS function in R

David Garibay, M.P.P.[*] Hawre Jalal, MD, Ph.D.[†]
Fernando Alarid-Escudero, Ph.D.[‡§]

#### Code function

This document presents an R implementation of the multivariate categorical sampling function mentioned in the “A Fast Nonparametric Sampling (NPS) Method for Time-to-Event in Individual-Level Simulation Models.” manuscript, and used in the R examples provided in the Appendix.

```
#' Nonparametric sampling of time to events from a discrete nonhomogeneous
#' Poisson point process
#'
#' \code{nps_nhppp} samples states for multiple individuals simultaneously.
#'
#' The elements of each row of the matrix `m_probs` should sum up to 1. In
#' case this does not happens, all the rows where this condition is not met
#' will be normalized.
#'
#' @param m_probs matrix with probabilities for each category. Each row
#' represents one individual and each column an specific category.
#' @param v_categories An optional argument. It is a vector containing the
#' name of the categories to sample from. The length of this vector must be
#' equal to the number of columns of `m_probs`.
#' @return A vector filled with sampled categories.
#' @export
nps_nhppp <- function(m_probs,
                      v_categories = NULL,
```

[*]School of Epidemiology and Public Health, Faculty of Medicine, University of Ottawa, Ottawa, ON, CA.
[†]School of Epidemiology and Public Health, Faculty of Medicine, University of Ottawa, Ottawa, ON, CA.
[‡]Department of Health Policy, Stanford University School of Medicine, Stanford, CA, USA.
[§]Center for Health Policy, Freeman Spogli Institute, Stanford University, Stanford, CA, USA.

```r
                     correction = c("none", "uniform")) {

  if (!is.numeric(m_probs)) {
    stop("`m_probs` must be a matrix filled with numeric elements")
  }

  if (!isTRUE(all.equal(target = rep(1, nrow(m_probs)),
                        current = as.numeric(rowSums(m_probs))))) {

    #* Find rows where the sum of their elements is not equal to 1.
    #* We use a very small value to check that they are equal to 1 since
    #* == and != sometimes do not work as desired. The value 1.5e-8
    #* was taken as the same level of tolerance as the `all.equal` function.
    v_rows_to_norm <- which(abs(1 - rowSums(m_probs)) > 1.5e-8)

    warning(
      "The rows: ",
      paste(as.character(v_rows_to_norm), collapse = ", "),
      " in `m_probs` do not sum up to 1. The values within these rows will be
      normalized and then used in the sampling process. In case this behaviour
      is not desired modify the content of `m_probs`.")

    # Normalize rows
    if (length(v_rows_to_norm) == 1) {
      m_probs[v_rows_to_norm, ] <- m_probs[v_rows_to_norm, ]/
        sum(m_probs[v_rows_to_norm, ])
    }
    if (length(v_rows_to_norm) > 1) {
      m_probs[v_rows_to_norm, ] <- m_probs[v_rows_to_norm, ]/
        rowSums(m_probs[v_rows_to_norm, ])
    }
  }

  # Number of categories to sample from
  n_cat <- ncol(m_probs)

  if (is.null(v_categories)) {
    #' Generate the numeric categories based on the number of columns of
    #' `m_probs`
    v_categories <- seq(0, (n_cat - 1))
  }
```

```
  correction <- match.arg(correction)

  # Number of time to events to draw
  n_samp <- nrow(m_probs)

  v_unif <- runif(n_samp, min = 0, max = 1)
  v_sum_p <- matrixStats::rowCumsums(m_probs)
  v_time_to_event <- v_categories[max.col(v_sum_p >= v_unif,
                                          ties.method = "first")]

  if (correction == "uniform") {
    v_time_to_event <- v_time_to_event + runif(n_samp, min = 0, max = 1)
  }
  return(v_time_to_event)

}
```

### Example 1 using python

#### Time to event from parametric hazards

David Garibay, M.P.P.[*] Hawre Jalal, MD, Ph.D.[†]
Fernando Alarid-Escudero, Ph.D.[‡§]

#### Code function

This document presents the python code corresponding to the first example presented in the "A Fast Nonparametric Sampling (NPS) Method for Time-to-Event in Individual-Level Simulation Models." manuscript. Since python has different parametrizations of the Gamma and Lognormal distributions, present in the example using R, this document only shows the code to replicate the example using the exponential function. .

```
# 01 Initial Setup ---------------------------------------------------------

# Import required modules
import numpy as np
import pandas as pd
import matplotlib.pyplot as plt
import scipy.stats as stats
import pandas as pd
```

```
# 02 General parameters -------------------------------------------------

# Exponential rate
rate = 0.1

# Sample size
n_samp = int(1e6)
```

[*]School of Epidemiology and Public Health, Faculty of Medicine, University of Ottawa, Ottawa, ON, CA.
[†]School of Epidemiology and Public Health, Faculty of Medicine, University of Ottawa, Ottawa, ON, CA.
[‡]Department of Health Policy, Stanford University School of Medicine, Stanford, CA, USA.
[§]Center for Health Policy, Freeman Spogli Institute, Stanford University, Stanford, CA, USA.

```
# 03 Data wrangling -----------------------------------------------------

# Obtain analytical values
a_true_mean   = 1/rate             ## mean
a_true_median = np.log(2)/rate   ## median
a_true_sd     = (1/(rate**2))**(1/2) ## SD

# Derive PDF from CDF
a_prob_exp_rates = (stats.expon.cdf(np.arange(1, 152), scale = 1/rate) -
                    stats.expon.cdf(np.arange(0, 151), scale = 1/rate))

# Normalize PDF
a_norm_exp_probs = a_prob_exp_rates/sum(a_prob_exp_rates)

# Sample values from normalized probabilites
a_random_exp_sample = np.random.choice(
  a       = np.arange(0, 151),
  size    = n_samp,
  replace = True,
  p       = a_norm_exp_probs)

# Add random number between 0 and 1 to approximate continuous time
a_random_exp_corr = (a_random_exp_sample +
                     np.random.random_sample(size = n_samp))
```

### Example 2 using python

#### Time to event from parametric hazards

David Garibay, M.P.P.[*] Hawre Jalal, MD, Ph.D.[†]
Fernando Alarid-Escudero, Ph.D.[‡§]

#### Code function

This document presents the python code corresponding to the second example presented in the "A Fast Nonparametric Sampling (NPS) Method for Time-to-Event in Individual-Level Simulation Models." manuscript.

```
# 01 Initial Setup ---------------------------------------------------------

# Import required modules
import numpy as np
import pandas as pd
import matplotlib.pyplot as plt
import scipy.stats as stats
import pandas as pd
```

```
# 02 Load base data -----------------------------------------------------

df_mort_data_raw = pd.read_csv(
  filepath_or_buffer = "../data/all_cause_mortality.csv")
```

[*]School of Epidemiology and Public Health, Faculty of Medicine, University of Ottawa, Ottawa, ON, CA.
[†]School of Epidemiology and Public Health, Faculty of Medicine, University of Ottawa, Ottawa, ON, CA.
[‡]Department of Health Policy, Stanford University School of Medicine, Stanford, CA, USA.
[§]Center for Health Policy, Freeman Spogli Institute, Stanford University, Stanford, CA, USA.

```
# 03 Data wrangling -----------------------------------------------------

# Keep data only for year 2015
df_mort_data_1 = df_mort_data_raw.query("Year == 2015").copy()

# Reset index (row ennumeration) of new data set
df_mort_data_1.reset_index(drop = True, inplace = True)

# Arrange data by sex, year and age values
df_mort_data_1.sort_values(by = ["Sex", "Year", "Age"], inplace = True)

# Group by sex
df_grouped = df_mort_data_1.groupby('Sex')

# Steps to derive the Instantaneous probability from the cumulative hazard
# H(t) - Cumulative hazard
df_mort_data_1['H_t'] = df_grouped['Rate'].cumsum()
# S(t) - Cumulative survival
df_mort_data_1['S_t'] = df_mort_data_1['H_t'].apply(lambda x: np.exp(-x))
# F(t) - Cumulative probability: 1 - S(t)
df_mort_data_1['F_t'] = 1 - df_mort_data_1['S_t']
# f(t) - Instantaneous probability
df_mort_data_1['p_t'] = (df_mort_data_1.groupby('Sex')['F_t'].
  diff().fillna(df_mort_data_1['F_t']))

# #* Check sum of probabilities
# df_mort_data_1.groupby(["Sex"])["p_t"].sum()

#* It is incomplete, so we will create a data frame to fill the probabilities
#* for each group
df_mort_append_0 = df_mort_data_1.groupby(["Sex"]).tail(1)

df_mort_append_0.loc[:, "Age"] = 101
df_mort_append_0.loc[:, "p_t"] = 1 - df_mort_append_0["F_t"]
df_mort_append_0.loc[:, "F_t"] = df_mort_append_0[["F_t", "p_t"]].sum(axis=1)
df_mort_append_0.loc[:, "S_t"] = 1 - df_mort_append_0["F_t"]
df_mort_append_0.loc[:, "H_t"] = np.nan
df_mort_append_0.loc[:, ["H_t", "Rate"]]  = np.nan


# Concatenate the original and the extra dataframes
df_mort_data_2 = pd.concat([df_mort_data_1, df_mort_append_0])
```

```
df_mort_data_2.sort_values(by = ["Sex", "Year", "Age"], inplace = True)
df_mort_data_2.reset_index(drop = True, inplace = True)

# # Now the sum of probs adds up to 1 for all groups
# df_mort_data_2.groupby(["Sex"])["p_t"].sum()

#* Convert into wide format
#* - Year will be discarded while turning data into wide format
df_mort_data_2_wide = df_mort_data_2.pivot(
  columns = "Age",
  index   = ["Year", "Sex"],
  values  = "p_t")

# Pass index into columns
df_mort_data_2_wide.reset_index(inplace = True)

# Remove row-axis name
df_mort_data_2_wide.rename_axis(None, axis = 1, inplace = True)
```

```
# 04 Sample times to events ---------------------------------------------
# We will use the "Total" Sex category to sample 100,000 individuals
a_age_values = np.arange(0, 102)
n_samples = int(1e5)

# Extract probability distribution for "Total" sex category
df_prob_values = (df_mort_data_2_wide.
  loc[df_mort_data_2_wide["Sex"] == "Total", 0:101])

# Convert into an array
a_prob_values = df_prob_values.iloc[0].to_numpy()

# # Plot probability distribution
# plt.rcParams["font.size"] = "22"
# plt.plot(a_age_values, a_prob_values, linewidth = 4)
# plt.title("Probability mass function of death by age in Total population,
#  2015")
# plt.grid(True, linestyle = "dotted", linewidth = 2)
# plt.show()
# plt.close()


# Instantiate base Data frame to fill with sampled ages of death
```

```
df_test_total = pd.DataFrame(
  data = {"Year": np.repeat(2015, n_samples), "Sex": "Total"})


# Set seed for reproducibility
np.random.seed(seed = 1234) # Set seed for reproducibility

#* Draw age of death from life tables using the `np.random.choice` function
df_test_total["age_death"] = np.random.choice(
  a       = a_age_values,
  size    = n_samples,
  replace = True,
  p       = a_prob_values)

#* Add random number between 0 and 1 to approximate continous time
df_test_total["age_death_corr"] = (df_test_total["age_death"] +
  np.random.random_sample(n_samples))

# Remove seed
np.random.seed(seed = None)
```

### Example 3 using python

**Time to event from parametric hazards**

David Garibay, M.P.P.[*] Hawre Jalal, MD, Ph.D.[†]
Fernando Alarid-Escudero, Ph.D.[‡§]

#### Code function

This document presents the python code corresponding to the third example presented in the "A Fast Nonparametric Sampling (NPS) Method for Time-to-Event in Individual-Level Simulation Models." manuscript.

```
# 01 Initial Setup ---------------------------------------------------------

# Import required modules
import numpy as np
import pandas as pd
import matplotlib.pyplot as plt
import scipy.stats as stats
import pandas as pd


# Define `nps_nhpp` function
def nps_nhpp(a_probs, correction, a_categories=None):

    valid_correction = {'none', 'uniform'}

    if correction not in valid_correction:
      print("Warning: incrrect inputs, allowed values: 'none' and 'uniform'")
      corresponding_values = None
```

[*]School of Epidemiology and Public Health, Faculty of Medicine, University of Ottawa, Ottawa, ON, CA.
[†]School of Epidemiology and Public Health, Faculty of Medicine, University of Ottawa, Ottawa, ON, CA.
[‡]Department of Health Policy, Stanford University School of Medicine, Stanford, CA, USA.
[§]Center for Health Policy, Freeman Spogli Institute, Stanford University, Stanford, CA, USA.

```
    if a_categories is None:
      # Get number of categories
      a_categories = np.arange(0, a_probs.shape[1])

    # Check that all PDF's sum up to 1
    if not all(np.isclose(a_probs.sum(axis = 1), 1)):
      a_probs = a_probs/a_probs.sum(axis=1, keepdims=True)

    # Get number of elements to draw
    a_samp_size = a_probs.shape[0]

    # Obtain array filled with random numbers following uniform distribution
    a_unif_probs = np.vstack(np.random.uniform(size = a_samp_size))

    # Get cumulative probabilities array from `a_probs`
    # Every row is a CDF
    a_cum_probs = np.cumsum(a_probs, axis = 1)

    # Compare uniform probabilities against cumulative probs
    comparison_result = a_cum_probs >= a_unif_probs

    # Getting positions where values are greater than or equal
    positions = np.argmax(comparison_result, axis = 1)

    corresponding_values = a_categories[positions]

    if correction == "uniform":
      corresponding_values = corresponding_values + a_unif_probs

    return corresponding_values
```

```
# 02 Load base data -----------------------------------------------------

df_mort_data_raw = pd.read_csv(
  filepath_or_buffer = "../data/all_cause_mortality.csv")
```

```
# 03 Data wrangling -----------------------------------------------------

# Keep data only for year 2015
df_mort_data_1 = df_mort_data_raw.query("Year == 2015").copy()
```

```
# Reset index (row ennumeration) of new data set
df_mort_data_1.reset_index(drop = True, inplace = True)

df_mort_data_1.sort_values(by = ["Sex", "Year", "Age"], inplace = True)

df_grouped = df_mort_data_1.groupby('Sex')

# H(t) - Cumulative hazard
df_mort_data_1['H_t'] = df_grouped['Rate'].cumsum()
# S(t) - Cumulative survival
df_mort_data_1['S_t'] = df_mort_data_1['H_t'].apply(lambda x: np.exp(-x))
# F(t) - Cumulative probability: 1 - S(t)
df_mort_data_1['F_t'] = 1 - df_mort_data_1['S_t']
# f(t) - Instantaneous probability
df_mort_data_1['p_t'] = (df_mort_data_1.groupby('Sex')['F_t'].
  diff().fillna(df_mort_data_1['F_t']))

#* Check sum of probabilities.
df_mort_data_1.groupby(["Sex"])["p_t"].sum()

#* It we incomplete, so we will create another row to fill the probabilities
df_mort_append_0 = df_mort_data_1.groupby(["Sex"]).tail(1)

df_mort_append_0.loc[:, "Age"] = 101
df_mort_append_0.loc[:, "p_t"] = 1 - df_mort_append_0["F_t"]
df_mort_append_0.loc[:, "F_t"] = df_mort_append_0[["F_t", "p_t"]].sum(axis=1)
df_mort_append_0.loc[:, "S_t"] = 1 - df_mort_append_0["F_t"]
df_mort_append_0.loc[:, "H_t"] = np.nan
df_mort_append_0.loc[:, ["H_t", "Rate"]]  = np.nan


# Concatenate the original and the extra dataframes
df_mort_data_2 = pd.concat([df_mort_data_1, df_mort_append_0])
df_mort_data_2.sort_values(by = ["Sex", "Year", "Age"], inplace = True)
df_mort_data_2.reset_index(drop = True, inplace = True)

# Now the sum of probs by Sex == 1
df_mort_data_2.groupby(["Sex"])["p_t"].sum()

#* Conver into wide format
#* - Year will be discarded while turning data into wide format
df_mort_data_2_wide = df_mort_data_2.pivot(
```

```
  columns = "Age",
  index   = ["Year", "Sex"],
  values  = "p_t")

df_mort_data_2_wide.reset_index(inplace = True)

df_mort_data_2_wide.rename_axis(None, axis = 1, inplace = True)
```

```
# 04 Sample times to events from heterogeneous cohorts ------------------

# Set seed for reproducibility
np.random.seed(seed = 1234)

# We will use the "Male" and "Female" sex categories
# to sample 100,000 individuals
a_sex = ["Male", "Female"]

# Sample size
n_samples = int(1e5)

# Allowed ages (0 to 102)
a_age_values = np.arange(0, 102)

# Sex proportions (50% males and 50% females)
p_sex = [0.5, 0.5]

# Instantiate base dataset
df_test_sex = pd.DataFrame(data = {"Year": np.repeat(2015, n_samples)})

# Draw sex of the individuals
df_test_sex["Sex"] = np.random.choice(
  a       = a_sex,
  size    = n_samples,
  replace = True,
  p       = p_sex)

# Append probability distribution based on Year and Sex
df_test_sex_probs = pd.merge(
  df_test_sex,
  df_mort_data_2_wide,
  how = "left",
  on  = ["Year", "Sex"])
```

```
# Obtain probability arrays
a_pob_probs = df_test_sex_probs.loc[:, 0:101].to_numpy()

#* Sample age of death for every individual distribution using the
#* `nps_nhpp` function
np.random.seed(seed = 234090) # Set seed for reproducibility
df_test_sex["age_death"] = nps_nhpp(
  a_probs    = a_pob_probs,
  correction = "none")

# Remove seed
np.random.seed(seed = None)
```

### Definition of the NPS function in python

David Garibay, M.P.P.[*] Hawre Jalal, MD, Ph.D.[†]
Fernando Alarid-Escudero, Ph.D.[‡§]

#### Code function

This document presents a python implementation of the multivariate categorical sampling function mentioned in the "A Fast Nonparametric Sampling (NPS) Method for Time-to-Event in Individual-Level Simulation Models." manuscript, and used in the R examples provided in the Appendix.

```
def nps_nhpp(a_probs, correction, a_categories=None):
    """
    Generate samples from different probability distributions using a
    multivariate categorical distribution.

    Parameters:
    -----------
    a_probs (array): Array filled with independent and equally-length
    probability distributions.
    correction (str): String defining whether to a random variable between 0
    and 1 to approximate continous time ("uniform") or no modification at all
    after performing the sample ("none").
    a_categories (array): (Optional) Array defining the names of the
    categories to sample.

    Returns:
    -----------
    numpy.array: Array filled with the sampled categories for all the
```

[*]School of Epidemiology and Public Health, Faculty of Medicine, University of Ottawa, Ottawa, ON, CA.
[†]School of Epidemiology and Public Health, Faculty of Medicine, University of Ottawa, Ottawa, ON, CA.
[‡]Department of Health Policy, Stanford University School of Medicine, Stanford, CA, USA.
[§]Center for Health Policy, Freeman Spogli Institute, Stanford University, Stanford, CA, USA.

```
probability distributions.

Example:
-----------
import numpy as np
import scipy.stats as stats

# Number of samples
n_samp = 100

# Create an array filled with categories between 1 and 101
a_categories = np.arange(1, 101)

# Parameters of normal distribution
## First distribution
norm_mean_1, norm_var_1 = 30, 10
## second distribution
norm_mean_2, norm_var_2 = 60, 10

# Get PDF from distributions
v_disc_PDF_norm_1 = stats.norm.pdf(a_categories, loc = norm_mean_1,
  scale = norm_var_1)
v_disc_PDF_norm_2 = stats.norm.pdf(a_categories, loc = norm_mean_2,
  scale = norm_var_2)

# normalize values
## First version
v_norm_PDF_norm_1 = v_disc_PDF_norm_1/sum(v_disc_PDF_norm_1)
## Second version
v_norm_PDF_norm_2 = v_disc_PDF_norm_2/sum(v_disc_PDF_norm_2)

# Join v_norm_PDF_norm_1 and v_norm_PDF_norm_2 into a sinlge array
a_PDF_1_2 = np.array([v_norm_PDF_norm_1, v_norm_PDF_norm_2])

# Randomly sample 0s and 1s
a_choice = np.random.choice(a = [0, 1], size = n_samp, replace = True,
  p = [0.5, 0.5])

# Use values present in a_choice to extract either v_norm_PDF_norm_1 or
# v_norm_PDF_norm_1 and stack them into an array
a_probs = np.stack(arrays = a_PDF_1_2[a_choice], axis = 0)
```

```
# Run the nps_nhpp function to sample times to events from the arrays
# of probabilities present in a_probs
nps_nhpp(a_probs= a_probs, correction="uniform")
-----------
"""
valid_correction = {'none', 'uniform'}

if correction not in valid_correction:
  print("Warning: correction argument only accepts: 'none' and uniform")

  corresponding_values = None

if a_categories is None:
  # Get number of categories
  a_categories = np.arange(0, a_probs.shape[1])

# Check that all PDF's sum up to 1
if not all(np.isclose(a_probs.sum(axis = 1), 1)):
  a_probs = a_probs/a_probs.sum(axis=1, keepdims=True)

# Get number of elements to draw
a_samp_size = a_probs.shape[0]

# Obtain array filled with random numbers following a uniform distribution
a_unif_probs = np.vstack(np.random.uniform(size = a_samp_size))

# Get cumulative probabilities array from `a_probs`
# Every row is a CDF
a_cum_probs = np.cumsum(a_probs, axis = 1)

# Compare uniform probabilities against cumulative probs
comparison_result = a_cum_probs >= a_unif_probs

# Getting positions where values are greater than or equal
positions = np.argmax(comparison_result, axis = 1)

corresponding_values = a_categories[positions]

if correction == "uniform":

  unif_corr = np.vstack(np.random.uniform(size = a_samp_size))
```

```
        corresponding_values = corresponding_values + unif_corr

    return corresponding_values
```
